# GENPHIRE: Enhancing Disease Risk Prediction Using Large Language Model

**DOI:** 10.64898/2025.12.03.25341576

**Authors:** Danwei Yao, Chang Liu, Shifan Yan, Jiayi Zhang, Yan V. Sun, Zhaohui S. Qin

## Abstract

**Background:** Estimating an individual’s liability to a disease is a fundamental problem in genome research. By exploiting findings from genome-wide association studies (GWASs), many powerful polygenic risk scores (PRSs) have been developed to predict disease risk based on genetic profile. Despite much success, the performance of PRS models is hindered by its inability to capture complex, nonlinear effects and interactions among variants.

**Results:** In this study, we introduce GENPHIRE or Genetic–Phenotypic Representation, a novel machine learning framework designed for disease risk prediction. The central idea in GENPHIRE is to translate an individual’s genotype profile to a “sentence” consist of basic clinical information together with an ordered list of top phenotypes for which the individual is found to have elevated number of risk alleles. After translation, the sentence is converted to an embedded vector by an pre-trained large language model (LLM) to assess its disease risk. We have tested GENPHIRE using UK Biobank data across a broad range of diseases and found it outperforms state-of-the-art PRS models more than 80% of the time.

**Conclusions:** Our results demonstrated that LLM-derived embeddings can be leveraged for disease risk prediction when an individual’s genotype profile is effectively represented. Our findings highlight a promising alternative strategy that complements existing PRS approaches.

## 1 Introduction

During the past two decades, the powerful genome-wide association study (GWAS) [1–4] has been successfully implemented in thousands of studies and identified a large number of single nucleotide polymorphisms (SNPs) that are significantly associated with a wide variety of complex traits [5, 6]. Despite exciting new discoveries of genetic associations, it becomes evident that the vast majority of these SNPs only have a small effect, meaning that each individual SNP only has limited predictive power [7, 8]. Nevertheless, in a landmark study, using a linear mixed model, Yang et al. demonstrated that much of the heritability of height can be explained by evaluating the effects of a large collection of SNPs simultaneously [7]. Since then, polygenic score (PGS) or polygenic risk score (PRS) (we will use PRS hereafter) has become the standard method in genetic-based disease risk prediction [9, 10] and played a critical role in precision genomic medicine [11]. PRS estimates an individual’s genetic predisposition to a disease of interest by comparing the person’s genotype profile with findings from matching GWASs [12]. Over the years, more and more sophisticated statistical methods such as linkage disequilibrium (LD) score regression [13] have been successfully applied to aggregated the effects of variants across the genome to predict phenotypes [8, 14–20]. More recently, functional annotations of SNPs derived from single cell omics essays have been incorporated to fine tune the weights used in PRS models to further improve their performance [21, 22]. In 2021, a dedicated and publicly available repository named PGS Catalog [23, 24] has been successfully established to compile and organize these models for broad research use.

Despite overwhelming success, most PRS models are linear models that do not take into account nonlinear effects and interactions, which hinder their performance in disease risk prediction. As a result, most PRS models only capture a limited proportion of disease variance. There is a pressing need for better methods to uncover genetic signals hidden in our genomes to enable more accurate disease risk prediction.

With the accumulation of big data, advanced machine learning methods such as deep neural networks have shown promising performance in many applications [25]. In particular, transformer-based pretrained large language models (LLM) [26] have demonstrated tremendous ability to solve a wide range of problems [27]. LLMs have already been enthusiastically embraced in biomedical research [28–30].

Recent advances in transformer-based large language models (LLMs) have demonstrated that pretrained models can capture deep semantic structure from biomedical text, enabling powerful new representations for downstream prediction tasks. GenePT, for example, leveraged GPT-3.5 to transform free-text NCBI gene descriptions into dense gene embeddings that improved performance across a range of functional genomics applications [31]. Likewise, a recent study showed that incorporating LLM-derived embeddings of electronic health record (EHR) concepts substantially enhanced early pancreatic cancer prediction, highlighting the broad utility of language models for biomedical risk stratification [32]. These findings suggest that LLMs can distill biologically meaningful context from domain vocabulary itself.

Inspired by these developments, we hypothesized that LLMs could also be used to encode genetic information by capturing the intrinsic relationships among phenotype traits linked to an individual’s genotype. Here, we introduce GENPHIRE (Genetic–Phenotypic Representation), a framework that transforms variant-level genetic data into phenotype-informed embeddings derived by LLM for disease risk prediction. The goal is to incorporate the complex interplay of an individual’s potential risks on a wide variety of phenotypes into the prediction model. We believe that semantics in the names of phenotypes with elevated potential risk can be captured by LLM and contributed to the prediction. This novel framework offers an attractive alternative and is complementary to mainstream PRS methodology.

## 2 Results

### 2.1 Overview of the framework

We developed GENPHIRE, a genetic–language embedding framework that transforms individual-level genotype data into dense, biologically-informed representations. Instead of constructing traditional PRS, GENPHIRE first maps each participant’s genetic profile into a phenotype space by aggregating risk-allele dosages associated with a comprehensive collection of 5,337 phenotypes selected from the GWAS Catalog [33]. These phenotype risk dosages are then normalized and ordered into a personalized genetic sentence, providing a compact and semantically structured summary of an individual’s genetic profile. We then leverage embeddings from pretrained LLMs to capture the latent relationships encoded in these genetic sentences. The resulting embeddings are used as input to a lightweight neural network classifier trained for binary disease prediction. For each disease, we trained a separate binary classifier using the genetic–language embeddings. An Overview of GENPHIRE is presented in Fig 1.

**Fig. 1.**
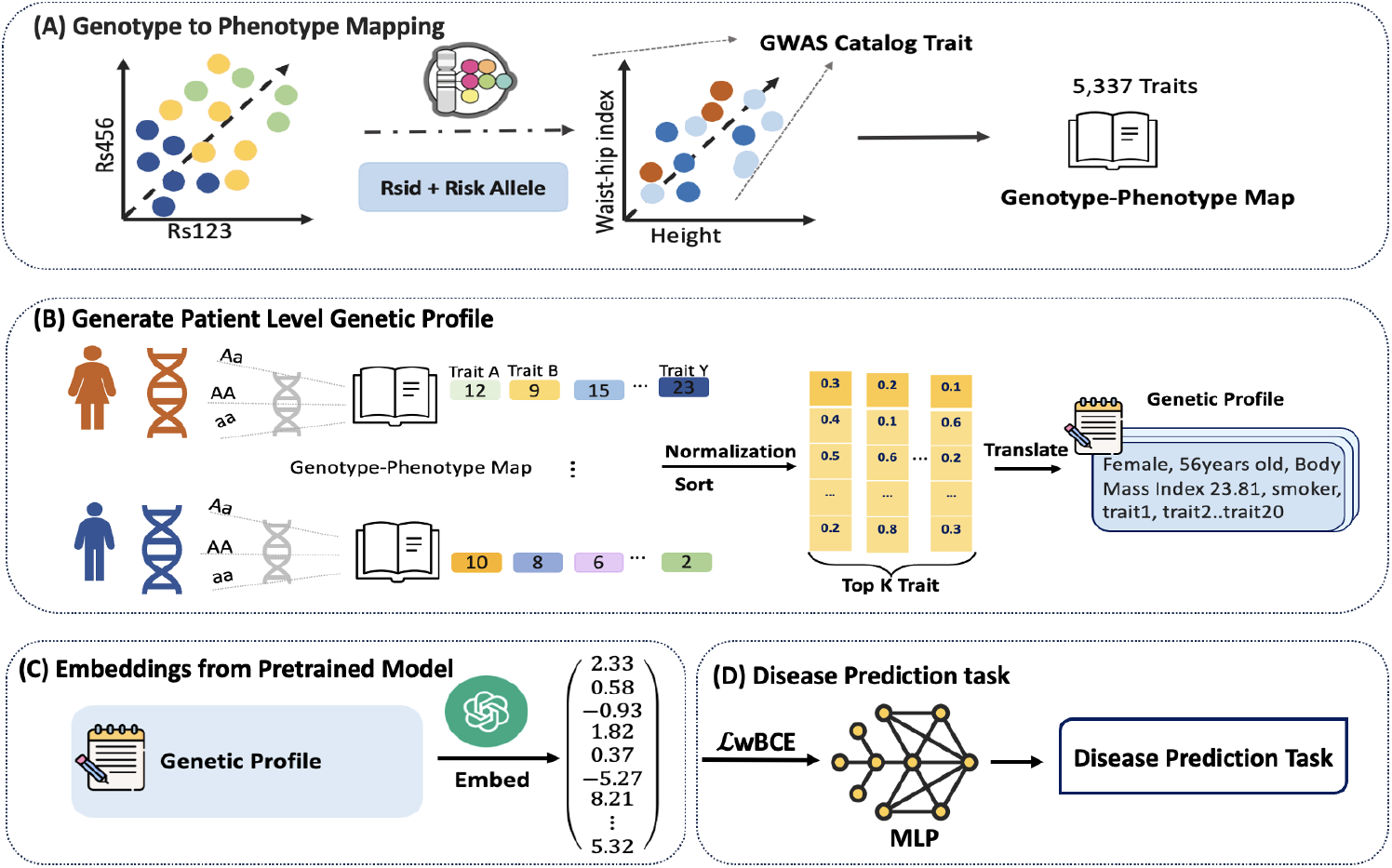
Overview of the GENPHIRE framework. (**A**) An individual’s genetic profile is first mapped to a phenotype space constructed using phenotypes stored in the GWAS catalog to create a genotype to phenotype mapping. (**B**) Individual-level genetic profiles are initially constructed by aggregating risk allele counts across traits. Following trait normalization and ranking, each patient’s profile is transformed into a structured “genetic sentence” that incorporates basic clinical information along with *K* top-ranked traits with the highest genetic risk burden. (**C**) Large language model (LLM)-derived embeddings are computed to encode semantic relationships among traits, forming patient-level embeddings. (**D**) Binary classification models are trained using the embeddings to predict disease risk.

### 2.2 Diseases studied

To evaluate GENPHIRE’s performance, we tested it on 21 different diseases selected to represent a wide range of disease conditions including cardiometabolic, vascular, respiratory, renal, musculoskeletal, and autoimmune conditions. The inclusion criteria for these diseases are reasonable prevalence (≥ 0.5%) and there are well defined International Classification of Diseases (ICD)-10 codes mapping to them.

All the case and control samples are selected from the UK Biobank (UKB), a large population-based cohort comprising over 500,000 participants aged 40–69 at baseline [34]. Following the literature, disease phenotypes were defined using ICD-10 codes. To be specific, disease status was defined using the UKB centrally derived “first occurrence” fields, based on any disease code mapped to 3-character ICD-10 categories. These variables integrate diagnosis information from multiple linked data sources, including hospital inpatient admissions, primary care records, national death registries (primary and secondary causes of death), and self-reported medical conditions.

For binary classification problem, whether the two classes have similar sample sizes is an important factor that influences the classification performance. To investigate the impact of imbalanced datasets on the performance of GENPHIRE, we considered four different ranges of imbalance ratio: 1:1 to 1:5; 1:5 to 1:15; 1:15 to 1:40; beyond 1:40. The corresponding prevalence ranges are: slightly imbalanced (prevalence greater than 16.6%), moderate imbalance (prevalence between 6.3% and 16.6%), severe imbalance (prevalence between 2.4% and 6.3%) and extreme imbalance (prevalence less than 2.4%). Here prevalence was defined as the presence of a recorded history of diagnosis based on these first occurrence fields. Participants with no qualifying ICD-10 code for a given condition were classified as controls. The complete list of the 21 diseases, along with their detailed definitions and corresponding ICD-10 code(s), is provided in Supplementary Table S1. The proportions of cases in the 21 diseases are presented in Supplementary Fig S1.

### 2.3 Performance evaluation

To evaluate the performance of GENPHIRE, we compared it against three popular PRS approaches, p-value thresholding (Pthres) [35], clumping+thresholding (C+T) (PRSice) [17, 36], and PRS-CS [19].

Five-fold cross-validation was used to evaluate the predictive performance of all the methods. Throughout the study, all model settings, including model architecture, class-weighting scheme, optimization hyperparameters, and train/test splits, were held constant across experiments to ensure a controlled comparison. Performance was evaluated using a variety of metrics including two types of area under curve (AUC) metrics: area under the receiver operating characteristic (AUROC) and area under the Precision Recall Curve (AUPRC), along with Mathews’ Correlation Coefficient (MCC) and F1 score. This comprehensive evaluation framework enables assessment of both overall discriminative ability and robustness at different levels of imbalance between cases and controls.

Overall, GENPHIRE demonstrated robust and broadly consistent performance. AUROC ranges from 0.57 to 0.78, with mean 0.71 and standard deviation 0.06 (Fig 2, complete results can be found in Supplementary Table S2). We found that AUROC ≥ 0.70 for the majority of the diseases (14 out of 21). The highest predictive performance was observed in Type 2 Diabetes (0.78), Diabetes (0.77), Heart Failure (0.76), Cardiovascular-Kidney-Metabolic (CKM) syndrome (0.76). For demonstration purpose, we present in Fig 3 results from four diseases: CKM syndrome, Chronic obstructive pulmonary disease (COPD), Myocardial infarction (MI), and Heart failure (HF).

**Fig. 2.**
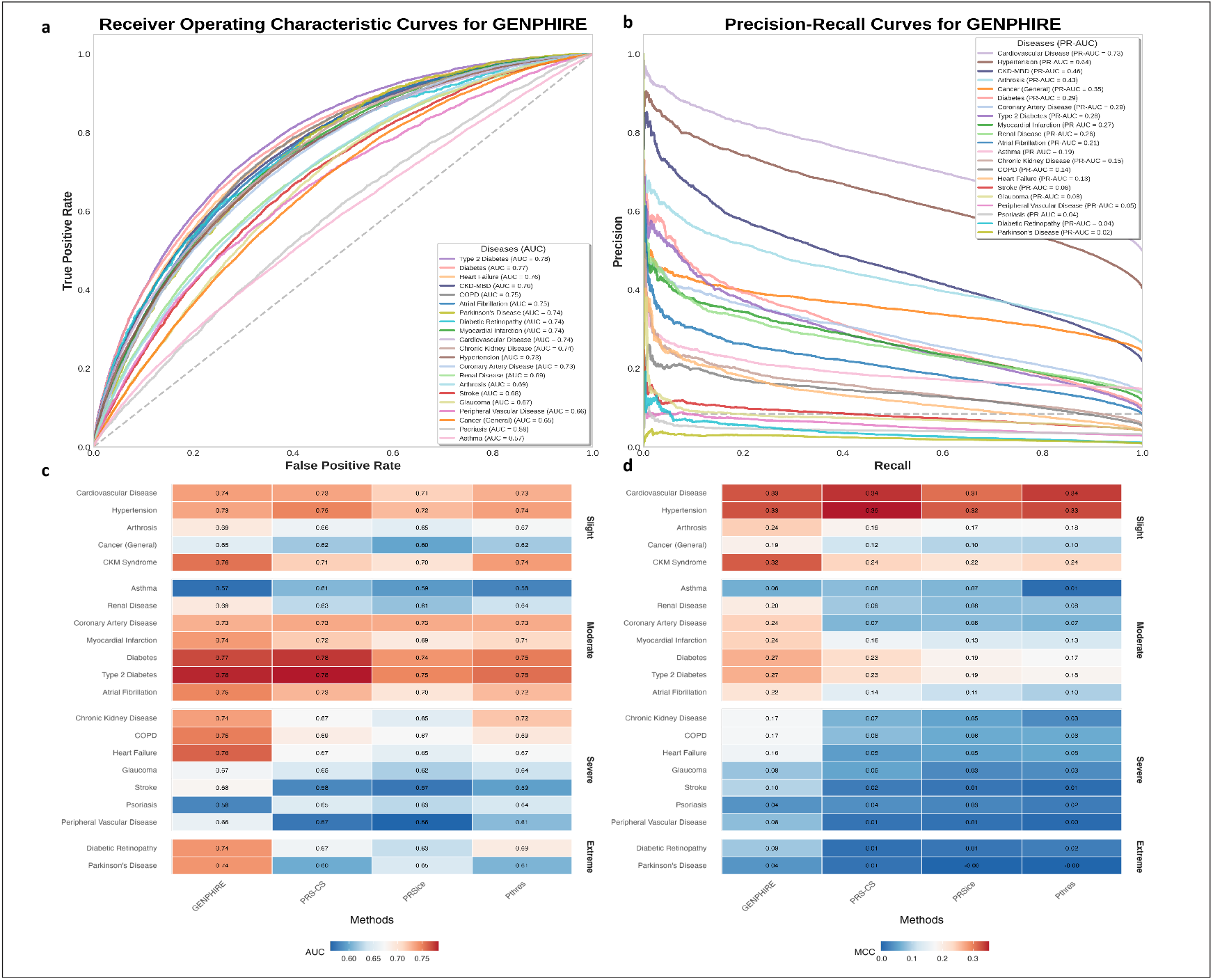
Performance comparison between GENPHIRE and polygenic risk score models across 21 diseases (a–b) Receiver operating characteristic (ROC) curve and Precision Recall curve (PRC) for GENPHIRE across 21 diseases. GENPHIRE demonstrates robust performance. **(c–d)** Comparison of GENPHIRE with three polygenic risk score (PRS) models (PR-CS, PRSice, Pthres) using area under ROC (AUROC) (panel c) and Matthews Correlation Coefficient (MCC; panel d). GENPHIRE matches or exceeds the best-performing PRS model in the majority of the 21 diseases. Among the PRS models, PRS-CS is the strongest competitor, outperforming GENPHIRE in four diseases by AUROC and in three diseases by MCC.

**Fig. 3.**
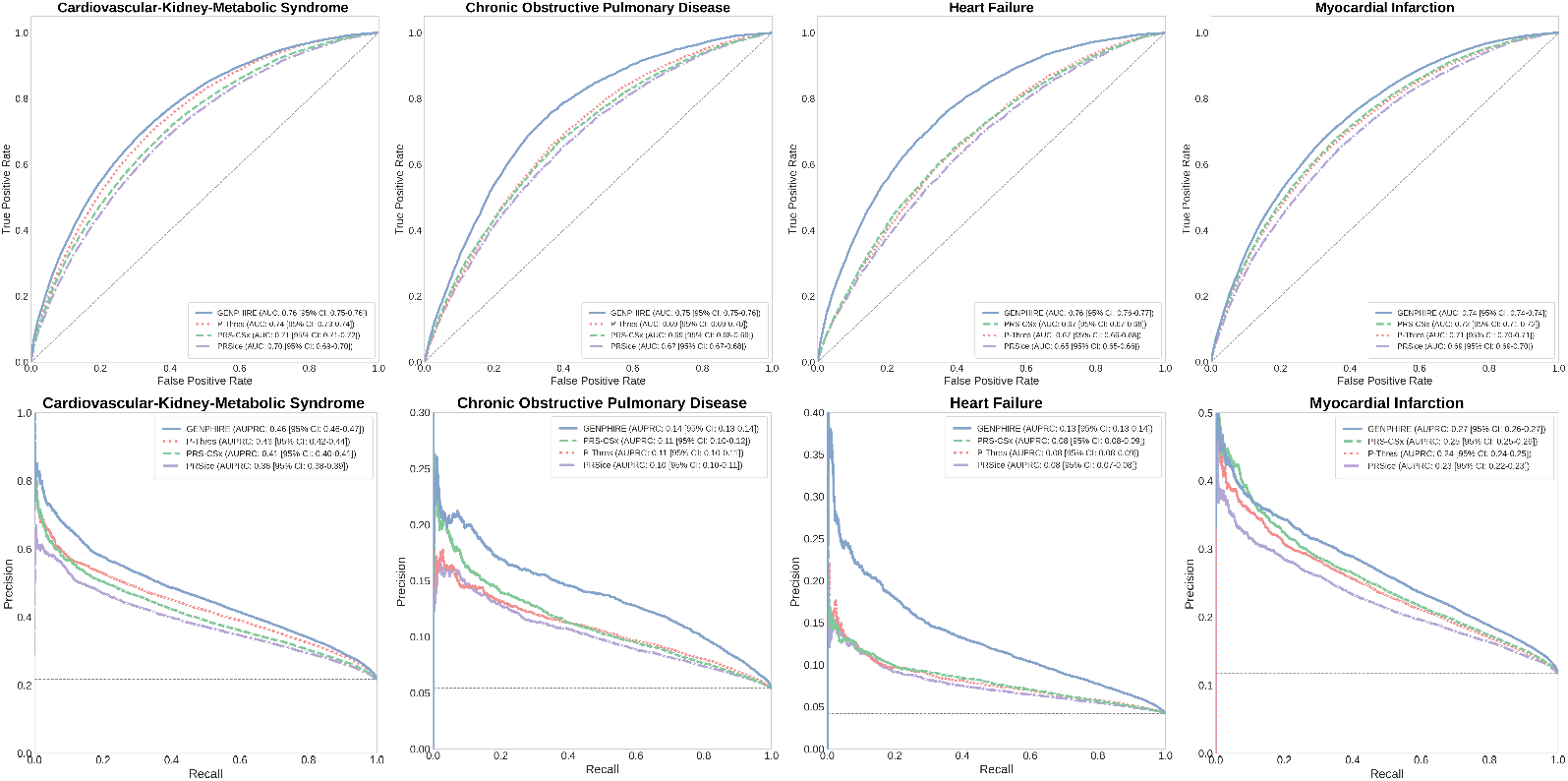
Predictive performance on four representative diseases. ROC curves (top row) and PRCs (bottom row) for four representative diseases—Cardiovascular–Kidney–Metabolic (CKM) syndrome, Chronic Obstructive Pulmonary Disease (COPD), Heart Failure (HF), and Myocardial Infarction (MI).

Next, we investigate whether the performance of GENPHIRE is affected by disease prevalence. We found that for diseases in the slightly imbalanced group, the model achieved high AUROC and AUPRC, indicating consistent predictive and descriptive performance aligned with the underlying prevalence distribution (Fig 2, Supplementary Table S2). Notably, for CKM syndrome, a composite and systemic condition involving intertwined metabolic disorders, chronic cardiovascular disease (CVD), and chronic kidney disease (CKD), our model yields strong performance (AUROC = 0.76; AUPRC = 0.46). We also examined two common cardiovascular-related diseases: COPD (prevalence 5.6%) and HF (prevalence 4.2%). For COPD, GENPHIRE achieved an AUROC of 0.75 (vs. 0.67–0.70 for PRS models) and an AUPRC of 0.14 (vs. 0.10–0.11). For Heart Failure, the model similarly outperformed all PRS models, achieving an AUROC of 0.76 (vs. 0.65–0.68) and an AUPRC of 0.13 (vs. 0.08–0.09). These case studies underscore the advantages of GENPHIRE in severely imbalanced, clinically complex diseases where conventional PRS approaches often struggle.

Compared to the three PRS models, we found that across all 21 diseases, GENPHIRE performs as well as or better than the best of the three PRS models in 17 diseases (81.0%), including most cardiometabolic, vascular, and renal diseases. The maximum improvement in AUROC is 18.9%. Among the three PRS-based models, PRS-CS consistently demonstrated to be the strongest overall performer, as expected given its incorporation of linkage disequilibrium structure and Bayesian effect-size shrinkage. PRS-CS outperforms GENPHIRE in four diseases (19.0%), with the most notable differences in Psoriasis (12.1% increase in AUROC); Asthma (7.0% increase in AUROC); Hypertension (2.7% increase in AUROC) and Diabetes (1.3% increase in AUROC).

When using MCC instead of AUROC, we found GENPHIRE achieved the highest or near-highest performance in 18 of 21 diseases (85.7%), whereas PRS-CS performs the best in three diseases: Asthma (by 33.3%), hypertension (by 6.1%), and CVD (by 3.0%). Overall, GENPHIRE delivers superior predictive accuracy for the majority of diseases we have tested, which is very encouraging.

### 2.4 Impact of case-control imbalance

For the purpose of comparing results across different levels of imbalance, we separate these 21 diseases into the aforementioned four groups: slightly imbalanced, moderate imbalanced, severe imbalanced and extreme imbalance, and produced four sets of side-by-side boxplots to compare performance (AUROC and MCC) for diseases belong to each of the four groups separately (Fig 4). We found that GENPHIRE outperforms all three PRS models across all four groups with notably stable results whereas the PRS models show progressively sharper deterioration as the imbalance problem worsens. And the margin of improvement of GENPHIRE becomes more pronounced in more imbalanced groups. These results demonstrate that GENPHIRE not only outperforms traditional PRS approaches for the majority of diseases we tested, but also remains more robust under increasing case–control imbalance. This is likely due to GENPHIRE’s conceptual simplicity and its ability to capture subtle genetic signals more effectively.

**Fig. 4.**
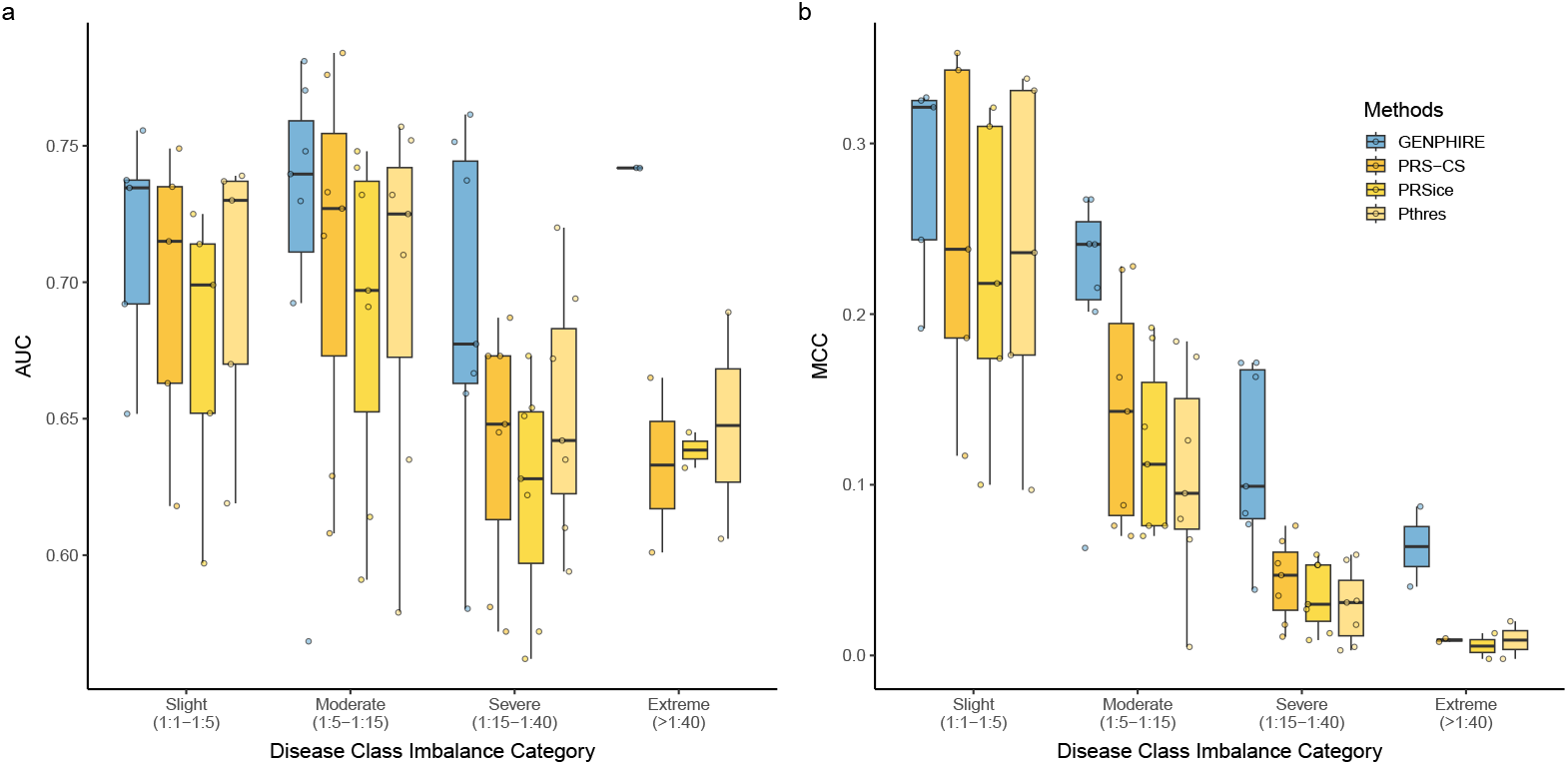
Performance comparison between GENPHIRE and polygenic risk score models grouped by the degree of case–control imbalance. Each boxplot summarizes the distribution of (a) AUROC and (b) MCC values for diseases within the Slight (1:1-1:5), Moderate (1:5-1:15), Severe (1:15-1:40), and Extreme (> 1 : 40) imbalance group. Blue denotes the proposed GENPHIRE framework, and the yellow palette corresponds to the three PRS models (PRS-CS, PRSice, and Pthres).

### 2.5 Performance in diverse population groups

It has been observed that PRS models show differential performance in diverse ethnic groups [37]. This has been a major barrier for the wide spread clinical implementation of the PRS models [38]. Therefore, it is of interest to evaluate how new disease risk prediction models such as GENPHIRE perform across different ethnic groups. To address this issue, we stratified UKB participants in the test set into four different ethnic groups–European, South Asian, African and others, and compared the performance of GENPHIRE with that of the three PRS models.

There is real but limited ethnic diversity in the UKB, with about 94.6% of all its participants were identified as white ethnicity [39]. The rest came from South Asian, African and other origin. In our test set, the breakdown of the four ethnic groups is: European (n=94,135, 93.9%), Others (n=2,865, 2.9%), South Asians(n=1,654, 1.6%), African (n=1,640, 1.6%). We plot four sets of side-by-side Violin plots, one for each method being tested, to show the difference in performance across ethnic groups (Fig 5).

**Fig. 5.**
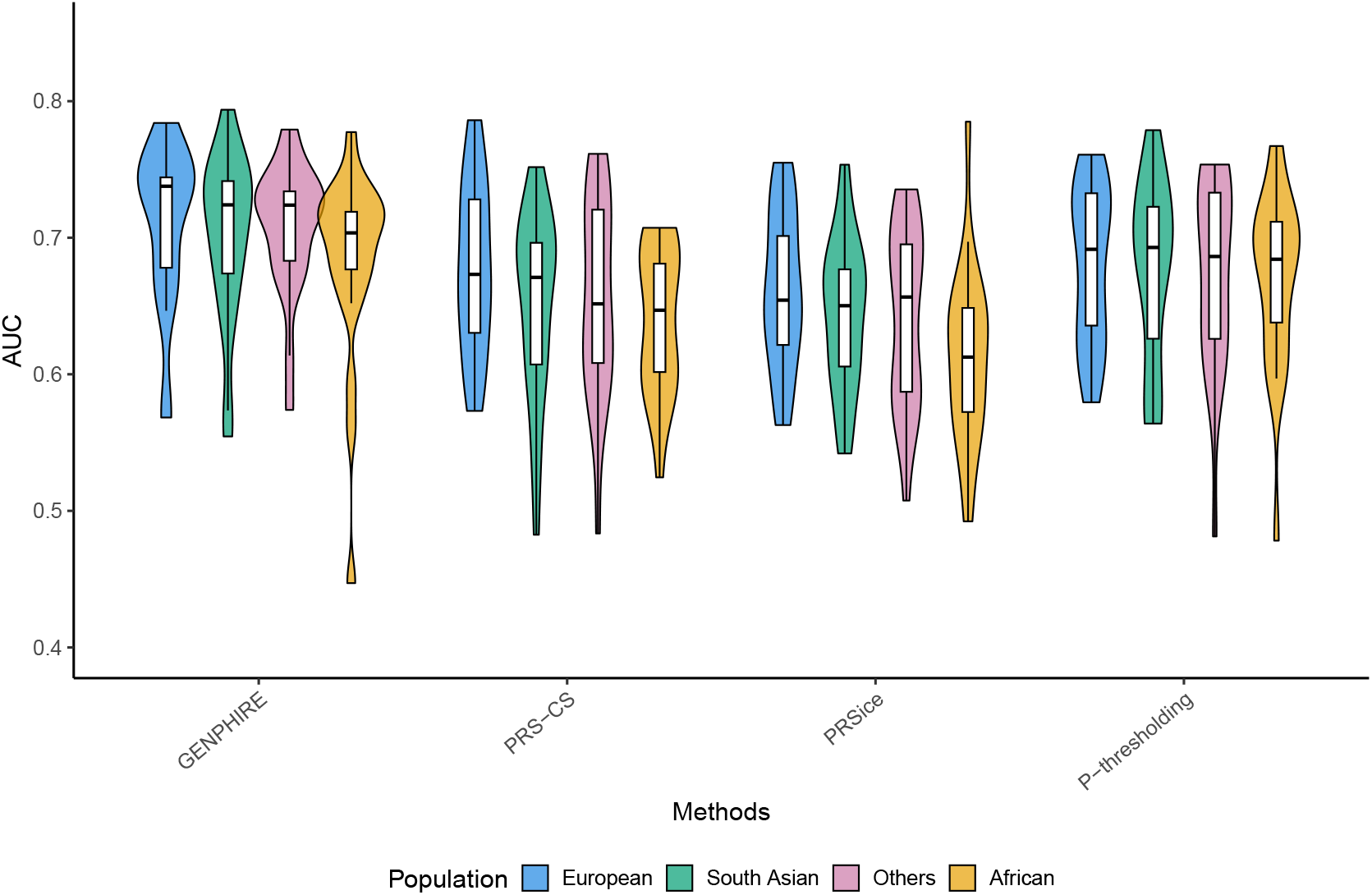
Performance comparison between GENPHIRE and polygenic risk score models across ethnic groups. Violin plots show AUROC distributions for GENPHIRE and three PRS models (PRS-CS, PRSice, Pthres) across four ethnic groups: European, South Asian, African, and Others. GENPHIRE performs consistently well across all four groups, with modest variation in AUROC. Europeans show the highest average AUROC (0.710; n=94,315), followed closely by Others (0.707, n=2,865) and South Asians (0.702; n=1,654). African-ancestry participants exhibit slightly lower performance (0.682; n=1,640). PRS models show slightly more variation across the ethnic groups, especially for PRS-CS and PRSice.

From these plots, we can see that GENPHIRE demonstrated consistent performance across the four ethnic groups which is very encouraging. Among the three PRS models, P-thres method is the most robust, similar to GENPHIRE with slightly more variation among ethnic groups. PRS-CS and PRSice exhibited a much greater degree of performance deterioration across the three non-European ethnic groups.

### 2.6 Model specifics

Beyond benchmarking against standard PRS methods, we are also interested in evaluating the influence of different pretrained embedding models and the number of top-ranked traits to understand their impact on predictive performance (complete result can be found in Supplementary Table S3).

To make an informed decision on which one to use as the embedding backbone within GENPHIRE, we generated patient-level embeddings using four different LLMs: OpenAI `text-3-small` (dimension=1,536): gemini-embedding-001 [40] (dimension =3,072), Voyage-3.5 (dimension=1,024), and the biomedical ontology–trained SapBERT encoder [41] (dimension=768). These models represent distinct families of text encoders: general-purpose LLMs and domain-specific biomedical transformers, enabling us to evaluate a wide variety of LLM options. For each embedding backbone, we applied the same genetic sentence construction procedure and trained separate classifiers for all 21 diseases. We found that the performance across the four embeddings are largely comparable, with SapBERT slightly underperform. Please see Supplementary Fig S2,S3. In the end, we decided to use the text-small embedding model from openAI as the backbone.

We also examined whether changing the number of top-ranked traits (*K*) affects GENPHIRE’S performance. As shown in Fig 6, when we chose *K* to be 15, 20, 25, 30, 50, the best overall AUROCs were achieved at *K* = 20 for most diseases: 0.770 (95% CI: 0.767–0.774) for diabetes, 0.735 (0.732–0.737) for HTN, 0.738 (0.733–0.742) for CKD, and 0.742 (0.731–0.754) for Parkinson’s disease. Similarly, the highest MCC values were also observed around *K* =20, reaching 0.327 (0.323–0.331) for HTN, 0.271 (0.261–0.273) for diabetes, 0.173 (0.167–0.179) for CKD, and 0.040 (0.031–0.048) for Parkinson’s disease (PD). Increasing *K* beyond 25 tended to reduce both AUROC and MCC, suggesting that adding lower-ranked traits introduces noise rather than a useful signal. As a result, we adopt *K* =20 as the default configuration for our framework.

**Fig. 6.**
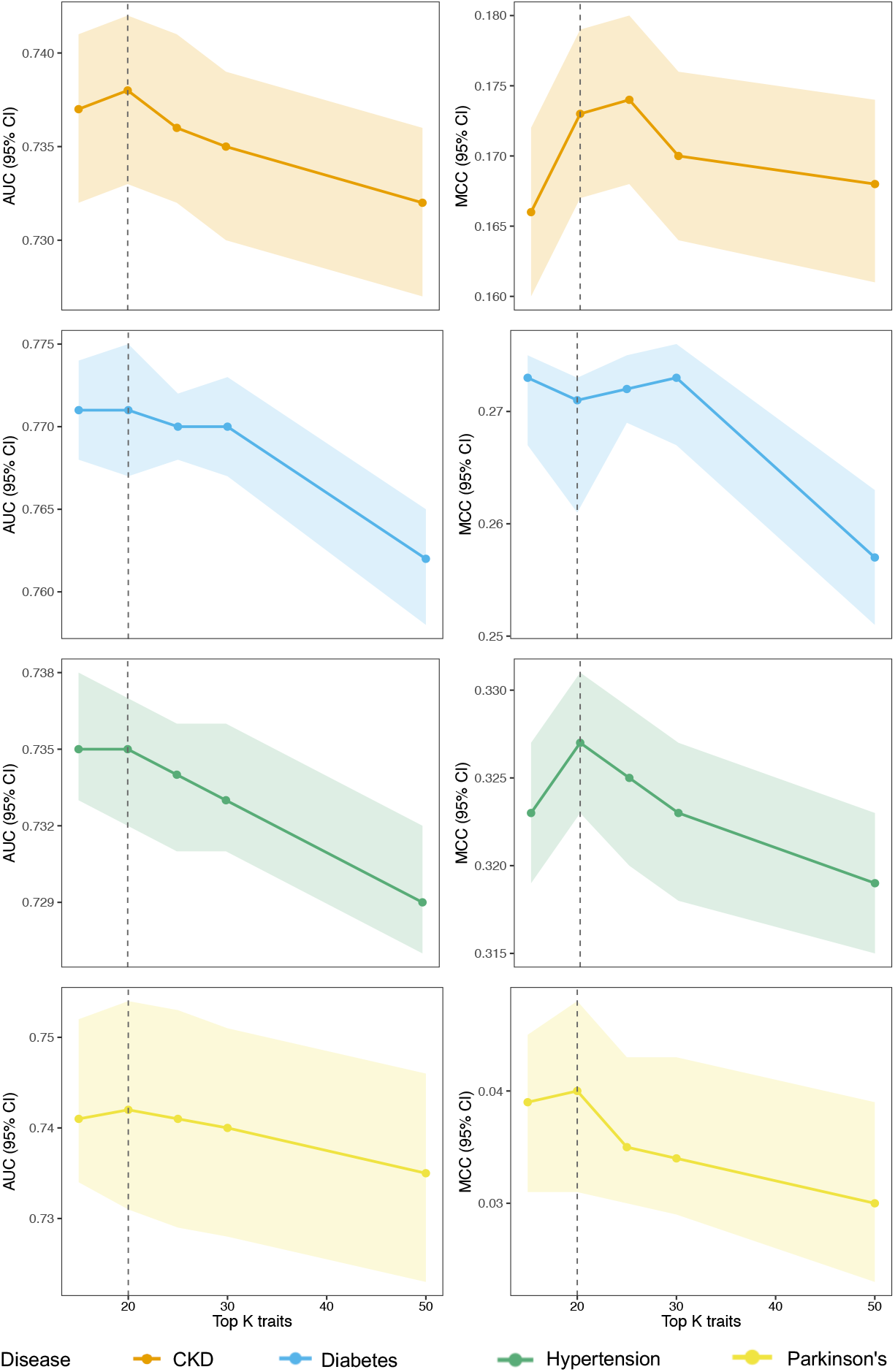
Performance comparison for GENPHIRE across varying different numbers of top-ranked traits for four representative diseases. Five different numbers of top-ranked traits was used: K = 15, 20, 25, 30, 50. Each panel shows mean AUC or MCC with 95% confidence intervals. Performance generally peaks or stabilizes around *K* ≈ 20, after which both AUC and MCC gradually decline.

We next conducted an ablation study to examine whether the order of the top 20 traits in the genetic profile sentence matters in the final performance. Across all evaluation metrics (AUROC, AUPRC, F1, and MCC), the full model consistently achieved the strongest performance. Removing trait ordering yielded a measurable but modest decline (change in AUROC: Δ_Hypertension_ = −0.020, Δ_Diabetes_ = −0.026, Δ_CKD_ = −0.026, Δ_Parkinson’s_ = −0.029), indicating that the ordering structure contributes meaningful information.

## 3 Discussion

In this study, we proposed GENPHIRE, a novel machine learning-based framework powered by LLM for disease risk prediction which is complementary to PRS methodology. To the best of our knowledge, this is the first attempt to apply LLMs to the disease risk prediction problem in human genetics.

Instead of fine-tuning weights of individual SNPs in PRS models, GENPHIRE translates the genotype profile into a short descriptive sentence. First, genetic risk from all major diseases and traits is summarized into a vector of risk allele counts, these counts are then normalized and sorted and the top 20 diseases or traits with the highest normalized genetic risk allele counts are identified. Next the names of these diseases, together with basic clinical information such as sex, age, BMI and smoking status, are concatenated together to form a short sentence to represent the genetic profile of the individual. Then LLM was utilized to classify these sentences into cases and controls. Testing on a broad range of complex diseases using UKB data demonstrated encouraging results. Across 21 different complex diseases we tested, GENPHIRE demonstrated better performance in the majority of them, especially on diseases with low to moderate prevalence.

Our findings are remarkable in three areas. First, GENPHIRE is a novel framework for estimating disease risk using genetic information and can be applied to nearly any disease, provided that a reasonably sized training dataset is available.

Second, GENPHIRE is conceptually simple. In contrast to applying more and more sophisticated strategies such as single cell epigenetics data [22] to fine tuning the weights of the PRS models, our strategy for calculating the genetic risk allele count is effectively unweighted PRS. And in the “genetic sentence”, only the ordered phenotypes names are used in the prediction model, rather than the actual risk-allele counts.

Third, GENPHIRE performs well and consistently regardless of the disease prevalence. This is important because the prevalence of many of the complex diseases is moderate at best. And we also observed that its performance is robust when applied to different ethnic groups, better than two out of three PRS models we tested.

A fundamental challenge with PRS models is that they require GWAS conducted on a large cohort to accurately estimate their model parameters. And conducting a large scale GWAS is extremely costly and time-consuming. For GENPHIRE, the genetic sentence can be derived in a distributed manner, like an encrypted individual-level genetic profile which make it possible to share among researchers without the risk of disclosing personal information of the participants. Therefore, it is possible to accumulate much larger training data to further improve GENPHIRE’s performance.

Admittedly, GENPHIRE has limitations and room for improvement. First, similar to other machine learning methods, given its deep neural network architecture, GENPHIRE requires a large amount of training data to achieve top-tier performance. Second, like many machine learning-based methods, GENPHIRE faced challenges with interpretability. Exactly how disease risk is estimated remains elusive at the current stage. Third, so far, GENPHIRE only takes in genotype data along with limited clinical information: BMI and smoking status. It would be interesting to add other covariates and environmental factors in the model so gene-environmental interaction may be incorporated.

In this study, we proposed a novel machine learning framework for individual-level disease risk prediction based on machine learning and LLM that demonstrated promising results. Compared to classical PRS models, we believed that the nonlinear and attention mechanisms of the neural network and LLM models give GENPHIRE advantages. The architecture of GENPHIRE supports the omnigenic [42] model proposed by Boyle et al. to characterize the underlying genetic mechanism influencing complex diseases. The “genetic sentence” representation technique we proposed is inspired by the “language-guided biology” idea proposed by Chen and Zou [31].

There are many different and powerful machine learning algorithms proposed in recent years. Many components used in GENPHIRE may not be ideal. Under the framework we proposed, we firmly believe that alternative representation methods and classification models exist that will deliver better performance than GENPHIRE.

## 4 Methods

### 4.1 Data

#### 4.1.1 UK Biobank genotype data

UKB [43] is a comprehensive cohort study that encompasses over 500,000 participants, age 40–69 years between 2006 and 2010. Our study included 502,366 participants who had genome-wide genetic data. According to UKB website, genotype calling was performed using arrays tailor-made by Affymetrix (now part of Thermo Fisher Scientific). There are two closely related custom arrays were used. 50,000 participants were run on the UK BiLEVE Axiom array and the remaining samples were run on the UK Biobank Axiom array. The final genotype dataset combines results from both arrays. There are 805,426 markers in the released genotype data. Quality control of SNP data and whole-genome SNP imputation was performed by the UKB analysis team [44].

#### 4.1.2 GWAS catalog

Established in 2008, The NHGRI-EBI GWAS catalog provides a consistent, searchable, visualizable and freely available database of SNP-trait associations [33]. The *All associations v1*.*0* dataset was downloaded from the GWAS Catalog on Jan 19 2025, which contains 569,163 published association records linking genetic variants to phenotypes. To avoid ambiguity in allele assignment, we restricted our analysis to single nucleotide polymorphisms (SNPs), which constitute the majority of genetic variants in the human genome. SNPs with unknown or ambiguous risk alleles were removed, resulting in a final set of 209,555 unique SNPs. Across these SNPs, the GWAS Catalog reports associations with 21,640 unique phenotypes (traits). While most phenotypes are linked to only a small number of SNPs, a few phenotypes (e.g., height) have hundreds or thousands of associated SNPs. In order to control variation due to the small sample size when calculating disease risk burden, we retained only phenotypes with more than five associated SNPs. This filtering yielded a total of 5,337 phenotypes for downstream analysis. Among these retained phenotypes, the median number of associated SNPs was 15 (interquartile range: 8–37; *X*_min_–*X*_max_: 6-16,707). The complete list of these 5,337 phenotypes can be found in Supplementary Table S4.

### 4.2 GENPHIRE method

The goal of GENPHIRE method is To obtain a concise yet informative genetic profile that can be understood by LLM, for which we developed the following embedding strategy which consists of the following four steps.

**Step 1** The input of the GENPHIRE algorithm is vectors of 805,426 dimension, which correspond to the number of SNPs genotyped by UKB. Each vector represents a UKB participant. In order to reduce the dimension of the genotype vector while maintaining as much functional information as possible, we first map the genotypes to the aforementioned 5,337 phenotypes. To be specific, for each individual, we calculated a phenotype-specific *genetic risk allele count* by aggregating the number of risk alleles across all SNPs associated with that phenotype in the GWAS Catalog. This is effectively an unweighted polygenic risk score. By doing this, we reduce the dimension more than 150 fold.

Let 𝒫= {*P*_1_, *P*_2_, …, *P*_*M*_ } denote the set of *M* phenotypes (diseases or traits) obtained from the GWAS Catalog, and let 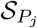 be the set of SNPs associated with phenotype *P*_*j*_. *M* = 5, 337 in this study.

Given an individual’s genotype for SNP 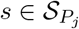, the *risk allele count g*_*s*_ was coded as:

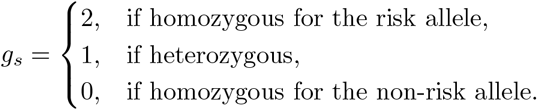

The genetic risk burden for phenotype *P*_*j*_ for individual *i* was then calculated as:

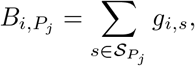

where *g*_*i,s*_ denotes the risk allele count of individual *i* at SNP *s. i* = 1, …*N, N* is the number of individuals. The complete genetic risk allele count vector for individual *i* is therefore:

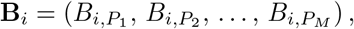

with each element representing the aggregated risk allele count for one phenotype. This produces a genetic profile that summarizes the cumulative effect of risk alleles in most known disease phenotypes.

**Step 2**. In this step, the goal is to further reduce the dimension of the genetic burden vector **B**_*i*_. We first normalized the risk allele counts for each trait across all individuals to obtain normalized genetic risk burden:

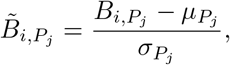

where 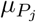 and 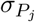 are the mean and standard deviation of risk allele counts for phenotype *P*_*j*_ across the cohort.

**Step 3**. For each individual *i*, we sorted the normalized genetic risk burden vector 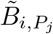 in descending order, and only retained the top *K* traits with the highest normalized genetic risk burden. In this study, we set *K* = 20.

**Step 4**. Subsequently, four clinical covariates—age (UKB field ID 21022), sex (UKB field ID 31), body mass index (BMI) (UKB field ID 21001), and smoking status (UKB field ID 20116) were concatenated to the top *k* trait names to form a structured short “genetic sentence” to characterize an individual’s genetic profile. For example:

> *“Male, 53 years old, BMI 22*.*6, non-smoker, Drinks per week, Waist circumference adjusted for body mass index, Glaucoma, Red cell distribution width, Triglycerides*, …*”*

**Step 5**. Finally, the constructed “genetic sentence” with approximately 50-100 words (median: 72 words; interquartile range: 68–76 words) was converted to a vector in an embedded space using LLM. In this study, we chose the embedding model text-embedding-ada-002 (with 1,536 dimension) provided by openAI to obtain embedding vectors:

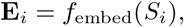

where *f*_embed_ : 𝒮 → ℝ^*d*^ maps sentences to a *d*-dimensional embedding space, *d* = 1,536.

The proposed GENPHIRE model is analogous to a multi-layer neural network model. The input is a genotype vector of 805,426 dimension, equal to the number of SNPs genotyped by UKB. In step 1 mentioned earlier, we convert the genotype vector into a vector of genetic risk burden with 5,337 dimensions. This is analogous to the convolution step in the convolutional neural network (CNN) [45]. In step 3, the 5,337-dimension genetic risk burden vector was sorted and converted to 20-dimension trait-names. This is analogous to a max pooling step in CNN. In step 4, the “genetic sentence” was mapped to a 1,536-dimension vector in the embedding space, which is achieved using a transformer-based neural network model.

### 4.3 Patient-level Genetic Language Embeddings

This sentence-to-embedding transformation allows us to represent an individual’s genetic and basic demographic profile as a dense representation suitable for downstream neural network–based disease risk prediction. The same genetic language embeddings were generated prior to any prediction task is carried out, or “pre-trained”, ensuring no target-specific information leakage.

### 4.4 Model Training and evaluation

#### Data split and leakage control

We partitioned the UK Biobank cohort of 502,366 participants into three mutually exclusive subsets using a 60%/20%/20% split: 301,420 for training, 100,473 for validation, and 100,473 for testing. Splits were *stratified* per disease to preserve case–control prevalence and were held constant across all experiments. All hyperparameter tuning and model selection used only the validation set; the test set was accessed *once* for final reporting.

#### Neural network models

We trained a separate binary classifier for each disease *d*, taking the individual embedding **E**_*i*_ (Section §Genetic Language Embedding) as input and produce a calibrated probability

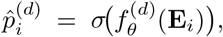

where 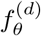 denotes a feed-forward neural network and *σ*(·) is the logistic sigmoid. We used a batch size of 512, the Adam optimizer with learning rate 1 × 10^−3^, and trained until the best validation performance was achieved (early stopping by checkpoint selection on validation AUROC). One model was fit per disease; no multi-task parameter sharing was used.

#### Class imbalance and objective

Given the imbalanced nature of many diseases, we optimized the *weighted* binary cross-entropy (wBCE) [46], with weights computed from the *training* split to avoid leakage. Let 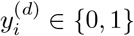 be the label for individual *i* and disease *d*, and let 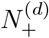 and 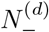 be the number of positive and negative training examples, respectively. We define class weights 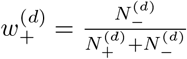 and 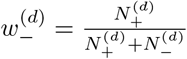. The per-disease training loss is

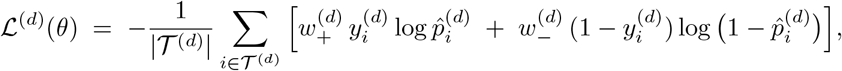

where 𝒯^(*d*)^ indexes the training set for disease *d*. This formulation is equivalent to `pos_ weight` reweighting in common deep learning libraries.

#### Primary and secondary metrics

The **primary** comparison metric was the area under the receiver operating characteristic curve (AUROC). As secondary metrics, we report the area under the precision–recall curve (AUPRC), the Matthews correlation coefficient (MCC), and the F1 score. The operating threshold *τ*^(*d*)^ for each disease was chosen on the *validation* set to maximize F1 score; these thresholds were then fixed and applied to the held-out test set. 95% CI for all metrics are estimated using non-parametric bootstrap resampling (1,000 replicates) of the held-out test set.

#### PRS model implementation

We first conducted GWAS on the *training* set which is the same one used to train GENPHIRE. The top 10 genotype principal components (PCs) were computed using PLINK [47], and age, sex, body mass index (BMI), and smoking status were included as covariates in the GWAS to control for ancestry and demographic effects. The resulting training-set summary statistics served as inputs for the three PRS models: (i) p-value thresholding (“Pthres”), (ii) clumping+thresholding (C+T; LD *r*^2^=0.1), and (iii) PRS-CS, a Bayesian shrinkage method with external LD reference.

For Pthres, we used PLINK 1.9 to compute scores under a grid of p-value thresholds, applying no LD pruning. We evaluated all thresholds on the validation set and selected the optimal threshold based on validation AUROC (test labels were held out) [48]. C+T was implemented via PRSice to perform clumping with a distance threshold of 250 kb and LD threshold of 0.1 [36]. The resulting clumped SNP set was filtered across a grid of p-value thresholds. Optimal threshold for each disease was determined using validation AUROC. For PRS-CS implementation, we used UKB European LD reference panel and ran the analysis with the automated optimization model parameter (phi) representing global shrinkage [19]. All PRS models and our embedding-based models were trained and evaluated with identical cohort splits and covariate (age, sex, BMI and smoking status) adjustments, ensuring a fair comparison.

### 4.5 Generalization study: cross-encoder robustness

To assess whether our genetic-language representation framework is robust to different embedding backbones, we generated patient-level embeddings using three additional models beyond OpenAI `text-3-small, dimension = 1,536`: gemini-embedding-001 (dimension = 3,072), Voyage-3.5 (dimension = 1,024), and the biomedical ontology–trained SAPBERT encoder (dimension=768). These models represent distinct families of text encoders: general-purpose LLMs and domain-specific biomedical transformers, allowing us to evaluate the architectural invariance of our framework.

For each embedding backbone, we applied the same genetic sentence construction procedure and trained separate classifiers for 21 diseases. All downstream settings, including model architecture, class-weighting scheme, optimization hyperparameters, and train/test splits, were held constant across experiments to ensure a controlled comparison. Performance on the fixed held-out test set was evaluated using AUROC, AUPRC, MCC, and F1 score.

This analysis quantifies the extent to which our framework is invariant to the choice of embedding model and evaluates whether general-purpose LLM embeddings or biomedical domain–specific embeddings produce more transferable patient representations across diverse disease outcomes.

### 4.6 Select optimal number of top-ranked traits

To evaluate whether changing the number of top-ranked traits (*K*) has any impact on GENPHIRE’s performance, we tested a series of different *K* (*K* = 15, 20, 25, 30 and 50) after normalization and ranking to construct the genetic sentences. Embeddings were generated following the same procedure described in Section *Patient-level Genetic Language Embeddings*. Four representative diseases were used for this evaluation: HTN, diabetes, CKD, and Parkinson’s disease, spanning slight, moderate, severe, and extreme imbalance settings. All models were trained using identical hyperparameter settings and the same text embedding backbone (OpenAI `text-3-small`) was applied to generate individual-level representations.

## 5 Data availability

The research was conducted using data from the UK Biobank Resource under application number 34031. The UKB study has approval from the North West Multicenter Research Ethics Committee as a Research Tissue Bank approval. For GWAS catalog traits information used for our analysis is provided at https://github.com/hereagain-Y/GENPHIRE.

## 6 Code availability

GENPHIRE is available at https://github.com/hereagain-Y/GENPHIRE.

## Acknowledgments

We thank members of the ENCORE group and Sun research group for helpful discussion.

## Supplementary Materials

**Table S1.** Definition of disease status in UK Biobank, including ICD–10 codes, UK Biobank first-occurrence fields, sample sizes (cases and controls), and prevalence. Diseases are ordered by prevalence in descending order, and grouped by four case-control imbalance groups.

| Phenotype | ICD-10 codes included | UK Biobank First-occurrence fields (Field IDs) | Controls, $n$ | Cases, $n$ | Prevalence (%) |
| --- | --- | --- | --- | --- | --- |
| **Slight Imbalance (1:1-1:5, Prevalence: $\geq 16.7\%$)** |  |  |  |  |  |
| Cardiovascular disease (CVD) | I00–I09, I11, I13, I20–I51, I10, I12, I15, I60–I69 | 131270–131356, 131286, 131290, 131294, 131360–131378 | 251 153 | 251 213 | 50.0 |
| Hypertension (HTN) | I10, I11, I12, I13, I15 | 131286, 131288, 131290, 131292, 131294 | 298 732 | 203 634 | 40.5 |
| Arthrosis | M15–M19 | 131868, 131870, 131872, 131874, 131876 | 368 099 | 134 267 | 26.7 |
| Cancer (any) | C00–C97 | 40005 | 378 595 | 123 771 | 24.6 |
| Cardiovascular-Kidney-Metabolic (CKM) syndrome | Composite ($\geq 2$ of: CAD/AFib/HF, T2D/obesity, HTN, CKD) | Derived from component conditions | 392 049 | 110 317 | 22.0 |
| **Moderate Imbalance (1:5-1:15, Prevalence: 6.3%–16.7%)** |  |  |  |  |  |
| Asthma | J45 | 131494 | 427 943 | 74 423 | 14.8 |
| Renal disease (broad) | N17–N23, N25–N29 | 132030–132052 | 432 994 | 69 372 | 13.8 |
| Coronary artery disease (CAD) | I20–I25 | 131296, 131298, 131300, 131302, 131304, 131306 | 435 108 | 67 258 | 13.4 |
| Myocardial infarction (MI) | I21–I23, I25 | 131298, 131300, 131302, 131306 | 443 701 | 58 665 | 11.7 |
| Obesity | E66 | 130792 | 449 282 | 53 084 | 10.6 |
| Diabetes (any) | E10–E14 | 130706, 130708, 130710, 130712, 130714 | 450 421 | 51 945 | 10.3 |
| Type 2 diabetes (T2D) | E11 | 130708 | 455 285 | 47 081 | 9.4 |
| Atrial fibrillation/flutter (AFib) | I48 | 131350 | 459 497 | 42 869 | 8.5 |
| **Severe Imbalance (1:15-1:40, Prevalence: 2.4%–6.3%)** |  |  |  |  |  |
| Chronic kidney disease (CKD) | N18 | 132032 | 471 130 | 31 236 | 6.2 |
| Chronic obstructive pulmonary disease (COPD) | J44 | 131492 | 474 176 | 28 190 | 5.6 |
| Heart failure (HF) | I50 | 131354 | 480 937 | 21 429 | 4.3 |
| Glaucoma | H40 | 131186 | 481 258 | 21 108 | 4.2 |
| Stroke | I60, I61, I63, I64 | 131360, 131362, 131366, 131368 | 481 351 | 21 015 | 4.2 |
| Psoriasis | L40 | 131742 | 486 368 | 15 998 | 3.2 |
| Peripheral vascular disease (PVD) | I73 | 131386 | 487 506 | 14 860 | 3.0 |
| **Extreme Imbalance ($> 1 : 40$, Prevalence $< 2.4\%$)** |  |  |  |  |  |
| Diabetic retinopathy | H36 | 131184 | 496 849 | 5 517 | 1.1 |
| Parkinson's disease | G20 | 131022 | 497 795 | 4 571 | 0.9 |

**Table S2.**
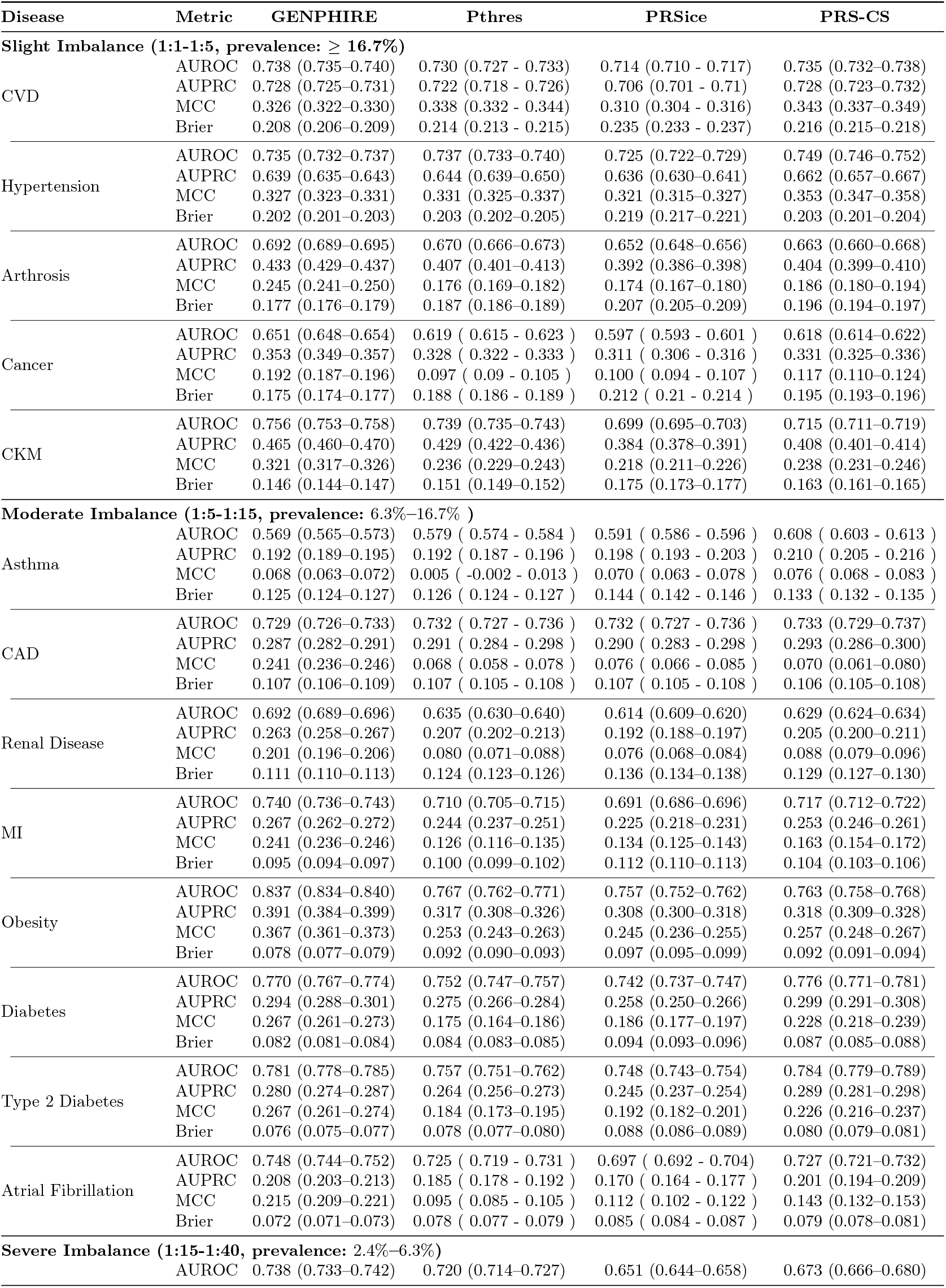
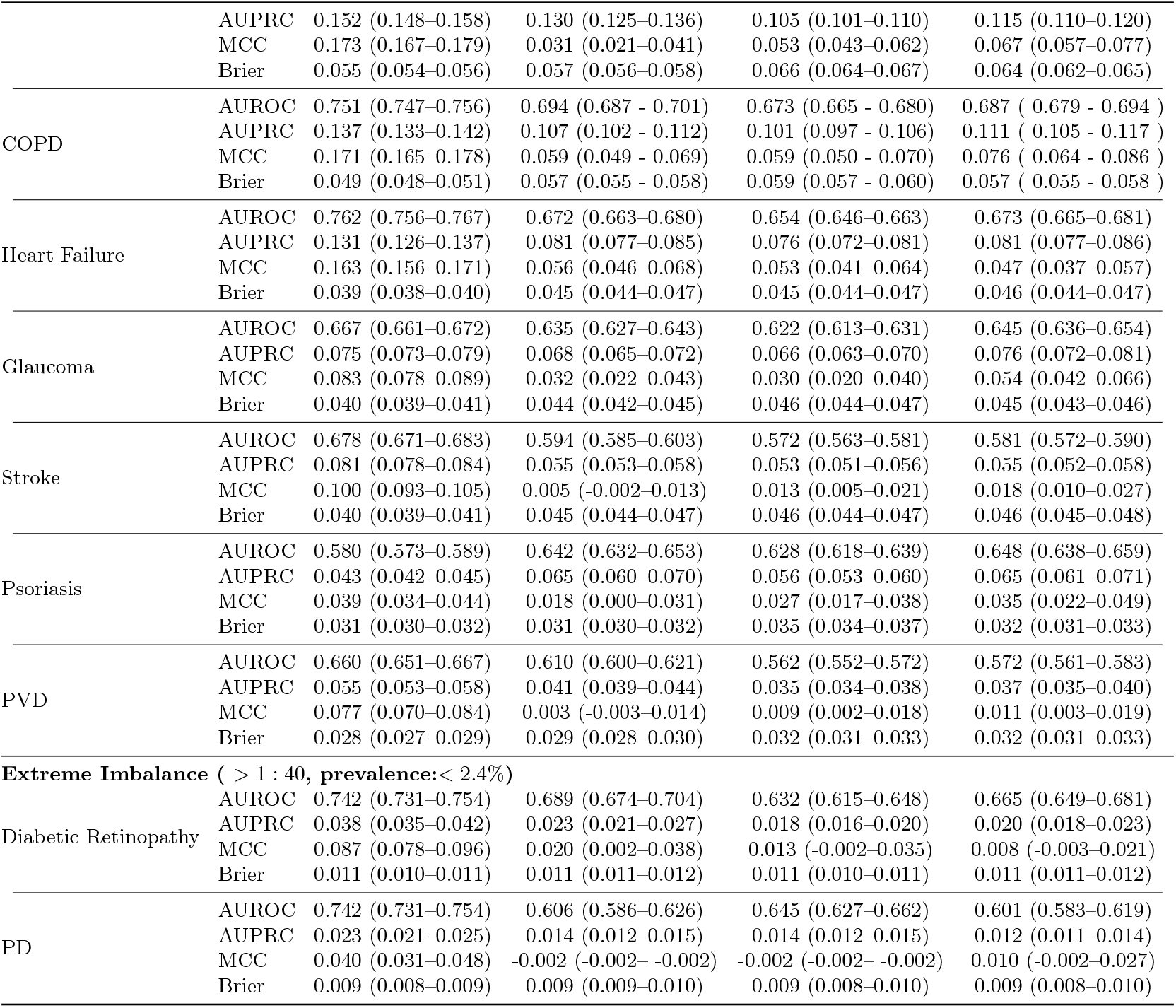
Prediction performance grouped by case-control imbalance level. Cells show mean (95% CI).

**Table S3.** Prediction performance grouped by case-control imbalance groups. Cells show mean (95% CI).

| Disease | Metric | OpenAI Embedding | Gemini Embedding | Voyage Embedding | SapBERT |
| --- | --- | --- | --- | --- | --- |
| **Slight Imbalance (1:1-1:5, prevalence: $\geq 16.7\%$)** |  |  |  |  |  |
| CVD | AUROC | 0.738 (0.735–0.740) | 0.737 (0.734-0.739) | 0.730 (0.727-0.732) | 0.732 (0.730–0.734) |
|  | AUPRC | 0.728 (0.725–0.731) | 0.726 (0.724-0.729) | 0.718 (0.715-0.720) | 0.722 (0.718–0.724) |
|  | MCC | 0.326 (0.322–0.330) | 0.321 (0.317-0.325) | 0.312 (0.308-0.316) | 0.317 (0.313–0.321) |
|  | F1 | 0.708 (0.707–0.710) | 0.708 (0.706-0.709) | 0.704 (0.703-0.706) | 0.706 (0.704–0.707) |
|  | Brier | 0.208 (0.207-0.208) | 0.208 (0.207-0.209) | 0.210 (0.210-0.211) | 0.209 (0.209–0.210) |
| Hypertension | AUROC | 0.735 (0.732–0.737) | 0.734 (0.731-0.736) | 0.726 (0.723-0.728) | 0.730 (0.728–0.733) |
|  | AUPRC | 0.639 (0.635–0.643) | 0.637 (0.633-0.640) | 0.625 (0.621-0.629) | 0.632 (0.628–0.635) |
|  | MCC | 0.327 (0.323–0.331) | 0.315 (0.311-0.319) | 0.315 (0.311–0.319) | 0.317 (0.312–0.321) |
|  | F1 | 0.640 (0.638–0.642) | 0.639 (0.636-0.641) | 0.637 (0.635–0.640) | 0.638 (0.635–0.640) |
|  | Brier | 0.208 (0.207-0.209) | 0.210 (0.209-0.211) | 0.212 (0.211–0.212) | 0.210 (0.210–0.211) |
| Arthrosis | AUROC | 0.692 (0.689-0.695) | 0.690 (0.688-0.693) | 0.685 (0.683-0.688) | 0.686 (0.683–0.689) |
|  | AUPRC | 0.433 (0.429–0.437) | 0.430 (0.426–0.434) | 0.424 (0.420–0.429) | 0.425 (0.421–0.429) |
|  | MCC | 0.245 (0.241–0.250) | 0.243 (0.239–0.248) | 0.236 (0.231–0.240) | 0.235 (0.230–0.239) |
|  | F1 | 0.489 (0.486–0.492) | 0.487 (0.484–0.490) | 0.484 (0.482–0.487) | 0.484 (0.481–0.486) |
|  | Brier | 0.227 (0.226–0.227) | 0.224 (0.224–0.225) | 0.225 (0.225–0.226) | 0.232 (0.231–0.232) |
| Cancer | AUROC | 0.651 (0.648–0.654) | 0.652 (0.649–0.655) | 0.650 (0.647–0.653) | 0.649 (0.646–0.653) |
|  | AUPRC | 0.353 (0.349–0.357) | 0.354 (0.350–0.357) | 0.352 (0.348–0.356) | 0.350 (0.346–0.354) |
|  | MCC | 0.192 (0.187–0.196) | 0.194 (0.189–0.198) | 0.190 (0.185–0.194) | 0.187 (0.182–0.191) |
|  | F1 | 0.442 (0.439–0.445) | 0.444 (0.441–0.447) | 0.442 (0.439–0.445) | 0.442 (0.439–0.444) |
|  | Brier | 0.229 (0.229–0.230) | 0.230 (0.230–0.231) | 0.230 (0.229–0.230) | 0.232 (0.232–0.233) |
| CKM | AUROC | 0.756 (0.753–0.758) | 0.756 (0.753–0.759) | 0.748 (0.745–0.750) | 0.740 (0.747–0.752) |
|  | AUPRC | 0.465 (0.460–0.470) | 0.464 (0.459–0.468) | 0.447 (0.442–0.452) | 0.450 (0.448–0.455) |
|  | MCC | 0.321 (0.317–0.326) | 0.321 (0.317–0.326) | 0.308 (0.304–0.313) | 0.313 (0.308–0.317) |
|  | F1 | 0.492 (0.489–0.496) | 0.492 (0.489–0.496) | 0.484 (0.481–0.487) | 0.487 (0.484–0.491) |
|  | Brier | 0.203 (0.202–0.204) | 0.204 (0.203–0.205) | 0.204 (0.203–0.205) | 0.207 (0.206–0.208) |
| **Moderate Imbalance (1:5-1:15, prevalence: 6.3%–16.7% )** |  |  |  |  |  |
| Asthma | AUROC | 0.569 (0.565–0.573) | 0.561 (0.557–0.565) | 0.558 (0.554–0.562) | 0.559 (0.555–0.563) |
|  | AUPRC | 0.192 (0.189–0.195) | 0.187 (0.184–0.190) | 0.182 (0.180–0.186) | 0.185 (0.182–0.188) |
|  | MCC | 0.068 (0.063–0.072) | 0.045 (0.041–0.050) | 0.052 (0.048–0.057) | 0.051 (0.046–0.056) |
|  | F1 | 0.267 (0.264–0.270) | 0.263 (0.260–0.265) | 0.262 (0.259–0.264) | 0.264 (0.262–0.267) |
|  | Brier | 0.250 (0.250–0.251) | 0.244 (0.244–0.245) | 0.250 (0.250–0.251) | 0.245 (0.245–0.246) |
| CAD | AUROC | 0.729 (0.726–0.733) | 0.729 (0.725–0.732) | 0.726 (0.723–0.730) | 0.722 (0.718–0.725) |
|  | AUPRC | 0.287 (0.282–0.291) | 0.287 (0.282–0.292) | 0.283 (0.278–0.288) | 0.279 (0.274–0.284) |
|  | MCC | 0.241 (0.236–0.246) | 0.239 (0.234–0.244) | 0.236 (0.231–0.242) | 0.230 (0.225–0.235) |
|  | F1 | 0.361 (0.357–0.365) | 0.359 (0.355–0.363) | 0.358 (0.353–0.362) | 0.353 (0.349–0.357) |
|  | Brier | 0.214 (0.213–0.215) | 0.217 (0.216–0.218) | 0.217 (0.216–0.218) | 0.218 (0.217–0.218) |
| Renal Disease | AUROC | 0.692(0.689-0.696) | 0.691 (0.688–0.695) | 0.687 (0.683–0.690) | 0.685 (0.681–0.688) |
|  | AUPRC | 0.263 (0.258–0.267) | 0.259 (0.254–0.264) | 0.253 (0.249–0.258) | 0.253 (0.248–0.257) |
|  | MCC | 0.201 (0.196–0.206) | 0.201 (0.195–0.206) | 0.194 (0.189–0.199) | 0.195 (0.190–0.200) |
|  | F1 | 0.333 (0.329–0.337) | 0.333 (0.328–0.337) | 0.328 (0.324–0.332) | 0.328 (0.325–0.332) |
|  | Brier | 0.218 (0.217–0.219) | 0.219 (0.218–0.219) | 0.228 (0.227–0.228) | 0.227 (0.226–0.227) |
| MI | AUROC | 0.740 (0.736–0.743) | 0.739 (0.736–0.742) | 0.736 (0.733–0.740) | 0.732 (0.729–0.736) |
|  | AUPRC | 0.267 (0.262–0.272) | 0.266 (0.261–0.271) | 0.262 (0.257–0.267) | 0.260 (0.255–0.265) |
|  | MCC | 0.241 (0.236–0.246) | 0.240 (0.234–0.245) | 0.238 (0.232–0.243) | 0.236 (0.230–0.241) |
|  | F1 | 0.344 (0.339–0.348) | 0.343 (0.338–0.348) | 0.341 (0.337–0.346) | 0.339 (0.335–0.343) |
|  | Brier | 0.211 (0.210–0.212) | 0.208 (0.207–0.209) | 0.216 (0.215–0.217) | 0.215 (0.214–0.216) |
| Obesity | AUROC | 0.837 (0.834–0.840) | 0.827 (0.824–0.829) | 0.824 (0.821–0.827) | 0.832 (0.830–0.835) |
|  | AUPRC | 0.391 (0.384–0.399) | 0.338 (0.333–0.345) | 0.335 (0.330–0.342) | 0.375 (0.368–0.382) |
|  | MCC | 0.367 (0.361–0.373) | 0.359 (0.354–0.365) | 0.358 (0.352–0.364) | 0.355 (0.349–0.360) |
|  | F1 | 0.439 (0.434–0.445) | 0.428 (0.423–0.433) | 0.428 (0.424–0.433) | 0.426 (0.421–0.431) |
|  | Brier | 0.174 (0.173–0.175) | 0.166 (0.165–0.167) | 0.181 (0.180–0.182) | 0.179 (0.178–0.180) |
| Diabetes | AUROC | 0.770 (0.767–0.774) | 0.765 (0.762–0.769) | 0.743 (0.739–0.746) | 0.759 (0.756–0.763) |
|  | AUPRC | 0.294 (0.288–0.301) | 0.284 (0.278–0.290) | 0.249 (0.244–0.254) | 0.273 (0.268–0.279) |
|  | MCC | 0.267 (0.261–0.273) | 0.263 (0.257–0.269) | 0.238 (0.233–0.243) | 0.254 (0.248–0.259) |
|  | F1 | 0.351 (0.346–0.356) | 0.348 (0.343–0.353) | 0.325 (0.321–0.330) | 0.340 (0.335–0.345) |
|  | Brier | 0.205 (0.204–0.205) | 0.203 (0.202–0.204) | 0.212 (0.211–0.213) | 0.205 (0.205–0.206) |

Table S3 – continued from previous page
| Disease | Metric | OpenAI Embedding | Gemini Embedding | Voyage Embedding | SapBERT |
| --- | --- | --- | --- | --- | --- |
| Type 2 Diabetes | AUROC | 0.781 (0.778-0.785) | 0.776 (0.772-0.779) | 0.754 (0.750-0.757) | 0.769 (0.766-0.773) |
|  | AUPRC | 0.280 (0.274-0.287) | 0.274 (0.267-0.280) | 0.241 (0.235-0.246) | 0.259 (0.254-0.265) |
|  | MCC | 0.267 (0.261-0.274) | 0.262 (0.256-0.268) | 0.238 (0.231-0.243) | 0.250 (0.244-0.256) |
|  | F1 | 0.343 (0.337-0.348) | 0.338 (0.333-0.344) | 0.316 (0.311-0.320) | 0.327 (0.322-0.332) |
|  | Brier | 0.189 (0.188-0.190) | 0.182 (0.181-0.183) | 0.200 (0.199-0.201) | 0.205 (0.205-0.206) |
| Atrial Fibrillation | AUROC | 0.748 (0.744-0.752) | 0.745 (0.741-0.749) | 0.742 (0.738-0.746) | 0.740 (0.736-0.744) |
|  | AUPRC | 0.208 (0.203-0.213) | 0.201 (0.196-0.206) | 0.199 (0.195-0.204) | 0.197 (0.193-0.202) |
|  | MCC | 0.215 (0.209-0.221) | 0.208 (0.202-0.214) | 0.203 (0.197-0.208) | 0.206 (0.199-0.210) |
|  | F1 | 0.288 (0.283-0.293) | 0.281 (0.277-0.286) | 0.278 (0.273-0.283) | 0.278 (0.273-0.282) |
|  | Brier | 0.207 (0.206-0.208) | 0.219 (0.218-0.220) | 0.212 (0.211-0.213) | 0.217 (0.216-0.218) |
| **Severe Imbalance (1:15-1:40, prevalence: 2.4%-6.3%)** |  |  |  |  |  |
| CKD | AUROC | 0.738 (0.733-0.742) | 0.735 (0.730-0.739) | 0.731 (0.726-0.735) | 0.728 (0.724-0.733) |
|  | AUPRC | 0.152 (0.148-0.158) | 0.149 (0.145-0.153) | 0.146 (0.142-0.151) | 0.144 (0.140-0.149) |
|  | MCC | 0.173 (0.167-0.179) | 0.175 (0.169-0.180) | 0.170 (0.165-0.176) | 0.165 (0.160-0.172) |
|  | F1 | 0.227 (0.222-0.232) | 0.221 (0.217-0.225) | 0.219 (0.214-0.223) | 0.426 (0.421-0.431) |
|  | Brier | 0.215 (0.214-0.216) | 0.210 (0.209-0.211) | 0.215 (0.214-0.216) | 0.179 (0.178-0.180) |
| COPD | AUROC | 0.751 (0.747-0.756) | 0.749 (0.745-0.754) | 0.746 (0.742-0.751) | 0.745 (0.740-0.750) |
|  | AUPRC | 0.137 (0.133-0.142) | 0.135 (0.130-0.139) | 0.131 (0.127-0.135) | 0.131 (0.127-0.135) |
|  | MCC | 0.171 (0.165-0.178) | 0.182 (0.176-0.187) | 0.173 (0.167-0.179) | 0.170 (0.164-0.176) |
|  | F1 | 0.215 (0.210-0.220) | 0.214 (0.210-0.219) | 0.212 (0.207-0.216) | 0.210 (0.205-0.214) |
|  | Brier | 0.212 (0.211-0.213) | 0.217 (0.216-0.218) | 0.207 (0.206-0.208) | 0.205 (0.204-0.206) |
| Heart Failure | AUROC | 0.762 (0.756-0.767) | 0.757 (0.752-0.763) | 0.754 (0.748-0.759) | 0.747 (0.742-0.753) |
|  | AUPRC | 0.131 (0.126-0.137) | 0.127 (0.122-0.132) | 0.122 (0.118-0.128) | 0.122 (0.117-0.127) |
|  | MCC | 0.163 (0.156-0.171) | 0.171 (0.164-0.178) | 0.155 (0.148-0.162) | 0.150 (0.142-0.157) |
|  | F1 | 0.200 (0.193-0.206) | 0.197 (0.192-0.202) | 0.191 (0.185-0.197) | 0.442 (0.439-0.444) |
|  | Brier | 0.201 (0.200-0.202) | 0.200 (0.199-0.201) | 0.221 (0.220-0.222) | 0.216 (0.215-0.216) |
| Glaucoma | AUROC | 0.667 (0.661-0.672) | 0.669 (0.664-0.675) | 0.666 (0.660-0.672) | 0.665 (0.659-0.670) |
|  | AUPRC | 0.075 (0.073-0.079) | 0.076 (0.073-0.079) | 0.074 (0.072-0.077) | 0.075 (0.073-0.078) |
|  | MCC | 0.083 (0.078-0.089) | 0.091 (0.086-0.096) | 0.078 (0.073-0.084) | 0.082 (0.077-0.088) |
|  | F1 | 0.126 (0.122-0.129) | 0.128 (0.125-0.132) | 0.123 (0.120-0.128) | 0.041 (0.034-0.048) |
|  | Brier | 0.232 (0.231-0.231) | 0.223 (0.223-0.224) | 0.220 (0.220-0.221) | 0.228 (0.228-0.229) |
| Stroke | AUROC | 0.678 (0.671-0.683) | 0.676 (0.670-0.682) | 0.674 (0.668-0.679) | 0.670 (0.664-0.675) |
|  | AUPRC | 0.081 (0.078-0.084) | 0.080 (0.078-0.083) | 0.079 (0.076-0.082) | 0.078 (0.075-0.080) |
|  | MCC | 0.100 (0.093-0.105) | 0.101 (0.095-0.107) | 0.102 (0.096-0.107) | 0.098 (0.093-0.104) |
|  | F1 | 0.141 (0.136-0.146) | 0.142 (0.137-0.146) | 0.141 (0.137-0.145) | 0.136 (0.132-0.140) |
|  | Brier | 0.220 (0.219-0.220) | 0.226 (0.225-0.227) | 0.225 (0.224-0.226) | 0.222 (0.221-0.222) |
| Psoriasis | AUROC | 0.580 (0.573-0.589) | 0.581 (0.574-0.589) | 0.577 (0.570-0.585) | 0.574 (0.567-0.581) |
|  | AUPRC | 0.043 (0.042-0.045) | 0.043 (0.041-0.045) | 0.043 (0.042-0.045) | 0.042 (0.041-0.044) |
|  | MCC | 0.039 (0.034-0.044) | 0.040 (0.034-0.045) | 0.035 (0.030-0.041) | 0.030 (0.025-0.035) |
|  | F1 | 0.078 (0.075-0.082) | 0.079 (0.076-0.082) | 0.076 (0.073-0.079) | 0.074 (0.070-0.076) |
|  | Brier | 0.254 (0.253-0.254) | 0.246 (0.246-0.246) | 0.254 (0.253-0.254) | 0.247 (0.247-0.248) |
| PVD | AUROC | 0.660 (0.651-0.667) | 0.655 (0.648-0.664) | 0.652 (0.644-0.660) | 0.650 (0.643-0.659) |
|  | AUPRC | 0.055 (0.053-0.058) | 0.054 (0.052-0.057) | 0.053 (0.051-0.056) | 0.052 (0.050-0.055) |
|  | MCC | 0.077 (0.070-0.084) | 0.074 (0.067-0.081) | 0.078 (0.072-0.085) | 0.077 (0.070-0.083) |
|  | F1 | 0.107 (0.101-0.113) | 0.105 (0.099-0.111) | 0.104 (0.099-0.109) | 0.104 (0.099-0.108) |
|  | Brier | 0.232 (0.232-0.233) | 0.230 (0.229-0.230) | 0.239 (0.238-0.240) | 0.230 (0.230-0.231) |
| **Extreme Imbalance (> 1 : 40, prevalence:< 2.4%)** |  |  |  |  |  |
| Diabetic Retinopathy | AUROC | 0.742 (0.731-0.754) | 0.741 (0.729-0.753) | 0.724 (0.712-0.736) | 0.723 (0.713-0.736) |
|  | AUPRC | 0.038 (0.035-0.042) | 0.038 (0.033-0.042) | 0.035 (0.031-0.040) | 0.032 (0.029-0.036) |
|  | MCC | 0.087 (0.078-0.096) | 0.072 (0.061-0.081) | 0.063 (0.053-0.072) | 0.064 (0.054-0.072) |
|  | F1 | 0.087 (0.080-0.095) | 0.079 (0.070-0.088) | 0.073 (0.064-0.082) | 0.071 (0.063-0.078) |
|  | Brier | 0.186 (0.185-0.187) | 0.188 (0.188-0.189) | 0.215 (0.214-0.216) | 0.209 (0.208-0.209) |
| PD | AUROC | 0.742 (0.731-0.754) | 0.746 (0.735-0.757) | 0.738 (0.726-0.750) | 0.733 (0.722-0.744) |
|  | AUPRC | 0.023 (0.021-0.025) | 0.023 (0.021-0.025) | 0.022 (0.020-0.024) | 0.021 (0.019-0.022) |
|  | MCC | 0.040 (0.031-0.048) | 0.057 (0.049-0.065) | 0.057 (0.050-0.064) | 0.032 (0.025-0.040) |
|  | F1 | 0.046 (0.039-0.052) | 0.052 (0.046-0.057) | 0.047 (0.042-0.051) | 0.041 (0.034-0.048) |
|  | Brier | 0.241 (0.240-0.241) | 0.213 (0.213-0.214) | 0.221 (0.220-0.222) | 0.220 (0.218-0.220) |

**Table S4.** Summary of all GWAS catalog phenotypes used in the present study. (Excel file)

**Fig. S1.**
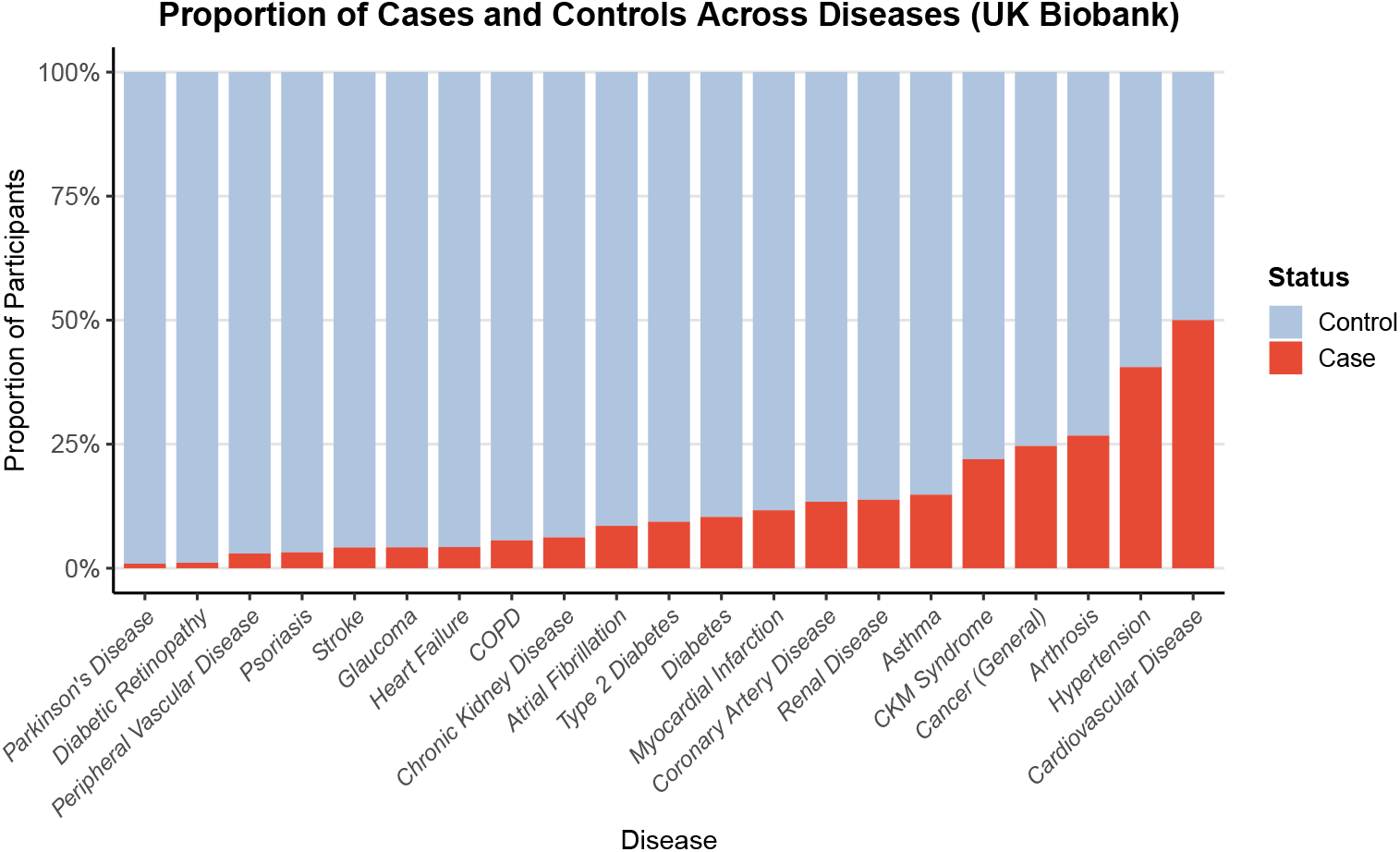
Proportions of cases and controls across 21 diseases in the UK Biobank. Stacked bar plots show the proportion of cases and controls in each of the 21 diseases analyzed.

**Fig. S2.**
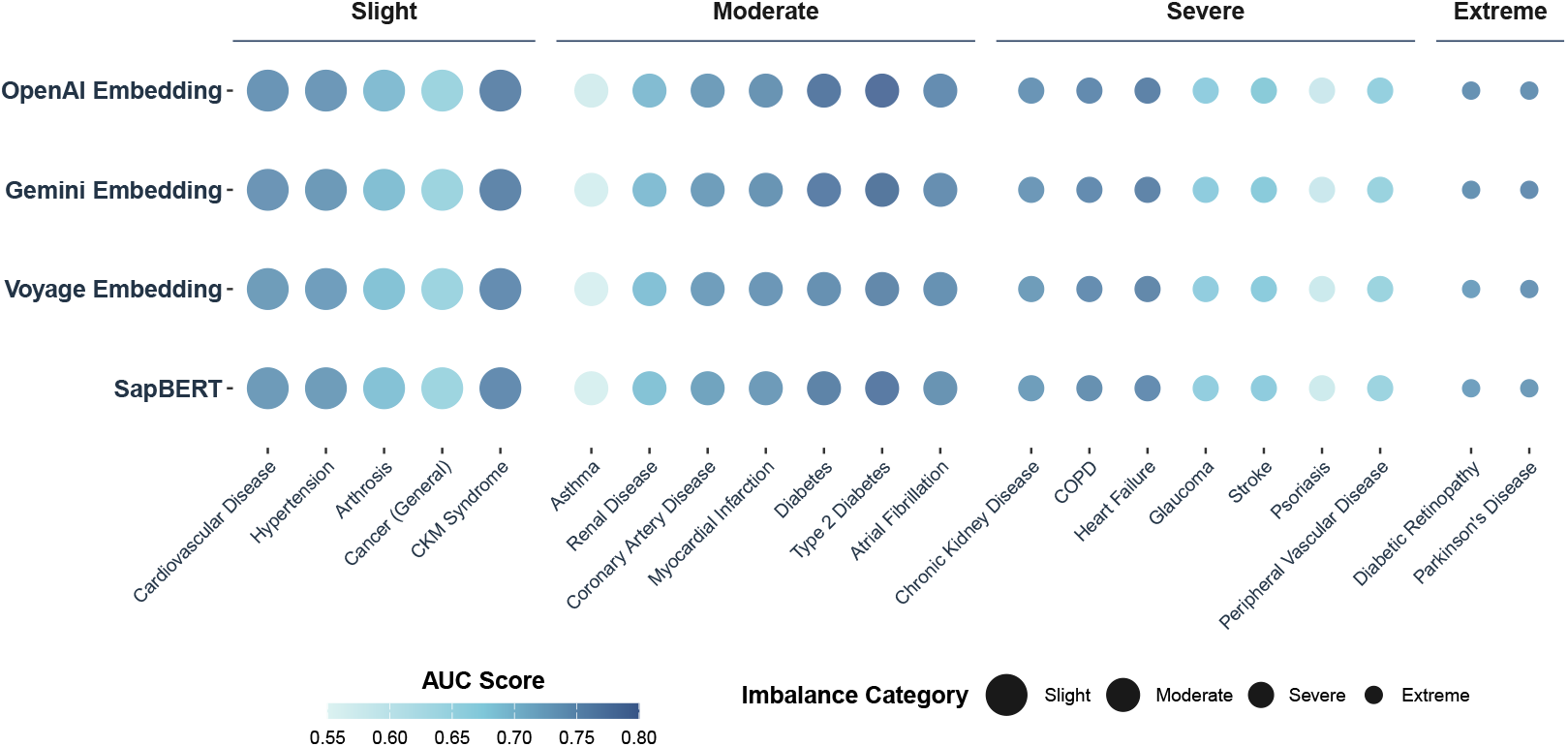
Performance comparison among embedding-based models across 21 diseases measured by AUROC. (slight imbalance → extreme imbalance). Bubble plots display AUROC values for four embedding models (OpenAI, Gemini, Voyage, SapBERT) evaluated across 21 diseases grouped by imbalance category. Bubble size reflects the imbalance level of each disease, while bubble color indicates AUROC performance. Across all imbalance groups, OpenAI, Gemini, and Voyage embeddings show consistently strong and stable performance, with higher AUROC values for most cardiometabolic and renal diseases. SapBERT performs competitively but shows slightly greater variability in the severe imbalance group. Overall, embedding-based genetic profile representations exhibit robust performance across a wide spectrum of class imbalance levels.

**Fig. S3.**
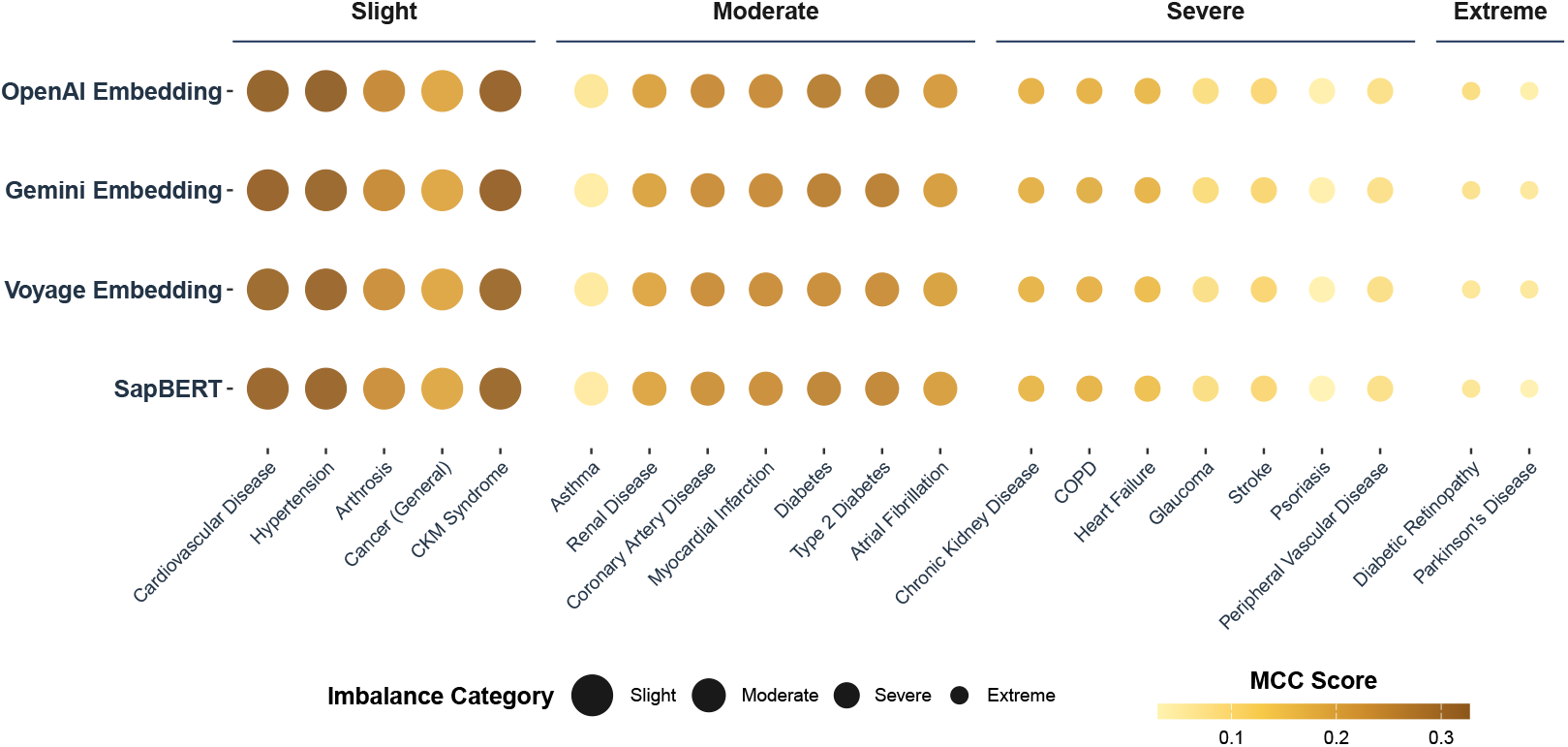
Performance comparison among embedding-based models across 21 diseases measured by MCC (slight imbalance → extreme imbalance). Bubble plots show Matthews Correlation Coefficient (MCC) scores for four embedding models (OpenAI, Gemini, Voyage, SapBERT) evaluated across 21 diseases grouped by imbalance category. Overall, embedding-based genetic profile representations exhibit robust performance across a wide spectrum of class imbalance levels.

## References

[1] Uffelmann, E., Huang, Q.Q., Munung, N.S., De Vries, J., Okada, Y., Martin, A.R., Martin, H.C., Lappalainen, T., Posthuma, D.: Genome-wide association studies. Nature Reviews Methods Primers 1(1), 59 (2021)

[2] Visscher, P.M., Brown, M.A., McCarthy, M.I., Yang, J.: Five years of gwas discovery. The American Journal of Human Genetics 90(1), 7–24 (2012)

[3] Visscher, P.M., Wray, N.R., Zhang, Q., Sklar, P., McCarthy, M.I., Brown, M.A., Yang, J.: 10 years of gwas discovery: biology, function, and translation. The American Journal of Human Genetics 101(1), 5–22 (2017)

[4] Abdellaoui, A., Yengo, L., Verweij, K.J., Visscher, P.M.: 15 years of gwas discovery: realizing the promise. The American Journal of Human Genetics 110(2), 179–194 (2023)

[5] Locke, A.E., Kahali, B., Berndt, S.I., Justice, A.E., Pers, T.H., Day, F.R., Powell, C., Vedantam, S., Buchkovich, M.L., Yang, J., et al.: Genetic studies of body mass index yield new insights for obesity biology. Nature 518(7538), 197–206 (2015)

[6] Pantelis, C., Papadimitriou, G.N., Papiol, S., Parkhomenko, E., Pato, M.T., Paunio, T., Pejovic-Milovancevic, M., Perkins, D.O., Pietiläinen, O., et al.: Biological insights from 108 schizophrenia-associated genetic loci. Nature 511(7510), 421–427 (2014)

[7] Yang, J., Benyamin, B., McEvoy, B.P., Gordon, S., Henders, A.K., Nyholt, D.R., Madden, P.A., Heath, A.C., Martin, N.G., Montgomery, G.W., et al.: Common snps explain a large proportion of the heritability for human height. Nature genetics 42(7), 565–569 (2010)

[8] Dudbridge, F.: Power and predictive accuracy of polygenic risk scores. PLoS genetics 9(3), 1003348 (2013)

[9] Lewis, C.M., Vassos, E.: Polygenic risk scores: from research tools to clinical instruments. Genome medicine 12(1), 44 (2020)

[10] Ding, Y., Hou, K., Xu, Z., Pimplaskar, A., Petter, E., Boulier, K., Privé, F., Vilhjálmsson, B.J., Olde Loohuis, L.M., Pasaniuc, B.: Polygenic scoring accuracy varies across the genetic ancestry continuum. Nature 618(7966), 774–781 (2023)

[11] Torkamani, A., Wineinger, N.E., Topol, E.J.: The personal and clinical utility of polygenic risk scores. Nature Reviews Genetics 19(9), 581–590 (2018)

[12] Lewis, C.M., Vassos, E.: Prospects for using risk scores in polygenic medicine. Genome medicine 9(1), 96 (2017)

[13] Bulik-Sullivan, B.K., Loh, P.-R., Finucane, H.K., Ripke, S., Yang, J., Patterson, N., 14 Daly, M.J., Price, A.L., Neale, B.M.: Ld score regression distinguishes confounding from polygenicity in genome-wide association studies. Nature genetics 47(3), 291–295 (2015)

[14] Palla, L., Dudbridge, F.: A fast method that uses polygenic scores to estimate the variance explained by genome-wide marker panels and the proportion of variants affecting a trait. The American Journal of Human Genetics 97(2), 250–259 (2015)

[15] Dudbridge, F.: Polygenic epidemiology. Genetic epidemiology 40(4), 268–272 (2016)

[16] Privé, F., Arbel, J., Vilhjálmsson, B.J.: Ldpred2: better, faster, stronger. Bioinformatics 36(22-23), 5424–5431 (2020)

[17] Choi, S.W., O’Reilly, P.F.: Prsice-2: Polygenic risk score software for biobank-scale data. Gigascience 8(7), 082 (2019)

[18] Mak, T.S.H., Porsch, R.M., Choi, S.W., Zhou, X., Sham, P.C.: Polygenic scores via penalized regression on summary statistics. Genetic epidemiology 41(6), 469–480 (2017)

[19] Ge, T., Chen, C.-Y., Ni, Y., Feng, Y.-C.A., Smoller, J.W.: Polygenic prediction via bayesian regression and continuous shrinkage priors. Nature communications 10(1), 1776 (2019)

[20] Zhou, G., Zhao, H.: A fast and robust bayesian nonparametric method for prediction of complex traits using summary statistics. PLoS genetics 17(7), 1009697 (2021)

[21] Amariuta, T., Ishigaki, K., Sugishita, H., Ohta, T., Koido, M., Dey, K.K., Matsuda, K., Murakami, Y., Price, A.L., Kawakami, E., et al.: Improving the trans-ancestry portability of polygenic risk scores by prioritizing variants in predicted cell-type-specific regulatory elements. Nature genetics 52(12), 1346–1354 (2020)

[22] Zhang, S., Shu, H., Zhou, J., Rubin-Sigler, J., Yang, X., Liu, Y., Cooper-Knock, J., Monte, E., Zhu, C., Tu, S., et al.: Single-cell polygenic risk scores dissect cellular and molecular heterogeneity of complex human diseases. Nature Biotechnology, 1–17 (2025)

[23] Lambert, S.A., Gil, L., Jupp, S., Ritchie, S.C., Xu, Y., Buniello, A., McMahon, A., Abraham, G., Chapman, M., Parkinson, H., et al.: The polygenic score catalog as an open database for reproducibility and systematic evaluation. Nature genetics 53(4), 420–425 (2021)

[24] Lambert, S.A., Wingfield, B., Gibson, J.T., Gil, L., Ramachandran, S., Yvon, F., Saverimuttu, S., Tinsley, E., Lewis, E., Ritchie, S.C., et al.: Enhancing the polygenic score catalog with tools for score calculation and ancestry normalization. Nature genetics 56(10), 1989–1994 (2024)

[25] Krizhevsky, A., Sutskever, I., Hinton, G.E.: Imagenet classification with deep convolutional neural networks. Advances in neural information processing systems 25 (2012)

[26] Zhao, W.X., Zhou, K., Li, J., Tang, T., Wang, X., Hou, Y., Min, Y., Zhang, B., Zhang, J., Dong, Z., et al.: A survey of large language models. arXiv preprint 2303.18223 1(2) (2023)

[27] Brown, T., Mann, B., Ryder, N., Subbiah, M., Kaplan, J.D., Dhariwal, P., Neelakantan, A., Shyam, P., Sastry, G., Askell, A., et al.: Language models are few-shot learners. Advances in neural information processing systems 33, 1877–1901 (2020)

[28] Simon, E., Swanson, K., Zou, J.: Language models for biological research: a primer. Nature Methods 21(8), 1422–1429 (2024)

[29] Nerella, S., Bandyopadhyay, S., Zhang, J., Contreras, M., Siegel, S., Bumin, A., Silva, B., Sena, J., Shickel, B., Bihorac, A., et al.: Transformers and large language models in healthcare: A review. Artificial intelligence in medicine 154, 102900 (2024)

[30] Hou, W., Ji, Z.: Assessing gpt-4 for cell type annotation in single-cell rna-seq analysis. Nature methods 21(8), 1462–1465 (2024)

[31] Chen, Y., Zou, J.: Simple and effective embedding model for single-cell biology built from chatgpt. Nature biomedical engineering 9(4), 483–493 (2025)

[32] Park, J., Patterson, J., Acitores Cortina, J.M., Gu, T., Hur, C., Tatonetti, N.: Enhancing ehr-based pancreatic cancer prediction with llm-derived embeddings. npj Digital Medicine 8(1), 465 (2025)

[33] Cerezo, M., Sollis, E., Ji, Y., Lewis, E., Abid, A., Bircan, K.O., Hall, P., Hayhurst, J., John, S., Mosaku, A., et al.: The nhgri-ebi gwas catalog: standards for reusability, sustainability and diversity. Nucleic acids research 53(D1), 998–1005 (2025)

[34] Collins, R.: What makes uk biobank special? The Lancet 379(9822), 1173–1174 (2012)

[35] Purcell, S., Wray, N., Stone, J., Visscher, P., O’Donovan, M., Sullivan, P.: Sklar, international schizophrenia consortium.(2009),” common polygenic vari ation contributes to risk of schizophrenia and bipolar disorder,”. Nature 460, 748–752 (2009)

[36] Euesden, J., Lewis, C.M., O’reilly, P.F.: Prsice: polygenic risk score software. Bioinformatics 31(9), 1466–1468 (2015)

[37] Kachuri, L., Chatterjee, N., Hirbo, J., Schaid, D.J., Martin, I., Kullo, I.J., Kenny, 16 E.E., Pasaniuc, B., Diverse Populations (PRIMED) Consortium Methods Working Group Auer Paul L. 20 Conomos Matthew P. 21 Conti David V. 22 23 Ding Yi 24 Wang Ying 19 25 26 Zhang Haoyu 27 28 Zhang Yuji 29, P.R.M., Witte, J.S., et al.: Principles and methods for transferring polygenic risk scores across global populations. Nature Reviews Genetics 25(1), 8–25 (2024)

[38] Martin, A.R., Kanai, M., Kamatani, Y., Okada, Y., Neale, B.M., Daly, M.J.: Clinical use of current polygenic risk scores may exacerbate health disparities. Nature genetics 51(4), 584–591 (2019)

[39] Fry, A., Littlejohns, T.J., Sudlow, C., Doherty, N., Adamska, L., Sprosen, T., Collins, R., Allen, N.E.: Comparison of sociodemographic and health-related characteristics of uk biobank participants with those of the general population. American journal of epidemiology 186(9), 1026–1034 (2017)

[40] Team, G., Anil, R., Borgeaud, S., Alayrac, J.-B., Yu, J., Soricut, R., Schalkwyk, J., Dai, A.M., Hauth, A., Millican, K., et al.: Gemini: a family of highly capable multimodal models. arXiv preprint 2312.11805 (2023)

[41] Liu, F., Shareghi, E., Meng, Z., Basaldella, M., Collier, N.: Self-alignment pretraining for biomedical entity representations. In: Proceedings of the 2021 Conference of the North American Chapter of the Association for Computational Linguistics: Human Language Technologies, pp. 4228–4238 (2021)

[42] Boyle, E.A., Li, Y.I., Pritchard, J.K.: An expanded view of complex traits: from polygenic to omnigenic. Cell 169(7), 1177–1186 (2017)

[43] Palmer, L.J.: Uk biobank: bank on it. The Lancet 369(9578), 1980–1982 (2007)

[44] Bycroft, C., Freeman, C., Petkova, D., Band, G., Elliott, L.T., Sharp, K., Motyer, A., Vukcevic, D., Delaneau, O., O’Connell, J., et al.: The uk biobank resource with deep phenotyping and genomic data. Nature 562(7726), 203–209 (2018)

[45] Krizhevsky, A., Sutskever, I., Hinton, G.E.: Imagenet classification with deep convolutional neural networks. Commun. ACM 60(6), 84–90 (2017)

[46] Ho, Y., Wookey, S.: The real-world-weight cross-entropy loss function: Modeling the costs of mislabeling. IEEE access 8, 4806–4813 (2019)

[47] Purcell, S., Neale, B., Todd-Brown, K., Thomas, L., Ferreira, M.A., Bender, D., Maller, J., Sklar, P., De Bakker, P.I., Daly, M.J., et al.: Plink: a tool set for whole-genome association and population-based linkage analyses. The American journal of human genetics 81(3), 559–575 (2007)

[48] Lin, Y.-P., Shi, Y., Zhang, R., Xue, X., Rao, S., Yin, L., Lui, K.F.H., Pan, D.J., Maurer, U., Choy, K.-W., et al.: A genome-wide association study of chinese and english language phenotypes in hong kong chinese children. npj Science of Learning 9(1), 26 (2024)

